# Machine Learning Enables Single-Score Assessment of MASLD Presence and Severity

**DOI:** 10.1101/2023.10.24.23297423

**Authors:** Robert Chen, Ben Omega Petrazzini, Girish Nadkarni, Ghislain Rocheleau, Meena Bansal, Ron Do

**Author notes:** Co-corresponding authors **Correspondence:** Meena Bansal, MD, Icahn Medical Institute, Floor 11 Room 70, 1425 Madison Avenue, New York, NY 10029, |, Ron Do, PhD, Annenberg Building, Floor 18 Room 80A, 1468 Madison Avenue, New York, NY 10029, |.

## Abstract

Metabolic dysfunction-associated steatotic liver disease (MASLD) affects 30% of the global population but is often underdiagnosed. To fill this diagnostic gap, we developed a digital score reflecting presence and severity of MASLD. We fitted a machine learning model to electronic health records from 37,212 UK Biobank participants with proton density fat fraction measurements and/or a MASLD diagnosis to generate a “MASLD score”. In holdout testing, our model achieved areas under the receiver-operating curve of 0.83-0.84 for MASLD diagnosis and 0.90-0.91 for identifying MASLD-associated advanced fibrosis. MASLD score was significantly associated with MASLD risk factors, progression to cirrhosis, and mortality. External testing in 252,725 diverse American participants demonstrated consistent results, and hepatologist chart review showed MASLD score identified probable MASLD underdiagnosis. The MASLD score could improve early diagnosis and intervention of chronic liver disease by providing a non-invasive, low-cost method for population-wide screening of MASLD.

## Introduction

Metabolic dysfunction-associated steatotic liver disease (MASLD), previously known as nonalcoholic fatty liver disease (NAFLD), is a preeminent cause of chronic liver disease on a global scale. Approximately 30% of the worldwide adult population is estimated to have MASLD;^1^ of whom as many as 40% will develop metabolic dysfunction-associated steatohepatitis (MASH).^2^ A small percentage will further progress to advanced fibrosis, the major risk factor for adverse liver outcomes.^3^

Despite its high prevalence, MASLD remains largely underdiagnosed due to its insidious onset; while limitations in current diagnostic practices hamper early intervention.^4^ Simple steatosis in early MASLD is largely asymptomatic, and although liver biopsy permits highly sensitive necroinflammation grading and fibrosis staging, it is costly and invasive. Non-invasive procedures like ultrasound, computerized tomography (CT), and magnetic resonance imaging (MRI) and their associated scores (e.g., FAST, MAST, MEFIB) can detect hepatic steatosis and/or fibrosis,^5–7^ but they have limitations in distinguishing between simple steatosis and MASH. Additionally, these procedures are inappropriate for large-scale population screening. Several laboratory-based scores are well suited for screening either the presence of MASLD or the degree of fibrosis;^8–10^ however, none accomplishes both tasks at once, potentially leading to unnecessary referrals and workup.

Addressing these issues, we aimed to develop a machine learning model that simultaneously predicts the presence and severity of MASLD, providing a holistic representation of a patient’s disease status. We used routine clinical and diagnostic data from 33,148 participants with proton density fat fraction (PDFF) measurements and 4,064 participants with diagnosed MASLD in the UK Biobank to construct the “MASLD score.” The score (a continuous spectrum from 0 to 3 with a higher score suggesting MASLD with more severe fibrosis) both captures the presence of MASLD and stratifies the degree of MASLD-associated fibrosis into F0-F2, intermediate, or F3-F4 categories based on the FIB-4 index. Because most patients with severe fibrosis do not have MASLD, we design the model to differentiate MASLD-associated fibrosis from fibrosis due to other causes. We assessed the model’s generalizability by testing on 252,725 participants from three diverse external cohorts: Mount Sinai Data Warehouse, Bio*Me* Biobank, and the *All of Us* Research Program. We also determined the model’s ability to identify underdiagnosis by performing chart review of 36 Bio*Me* participants without a MASLD diagnosis. Once integrated into electronic health record (EHR) systems,^11^ this model could use existing clinical data to automatically identify potential MASLD patients and recommend high-risk patients for gastroenterology referral and early intervention.

## Methods

### Data sources

We used data from four sources: UK Biobank, Mount Sinai Data Warehouse (MSDW), Bio*Me*, and *All of Us*. The UK Biobank includes 502,411 participants from across the United Kingdom; participants aged 40-69 were enrolled starting in 2006, and follow-up data was available for these participants until July 2022. The Mount Sinai Data Warehouse consists of Epic-derived electronic health records (EHRs) for more than 11 million patients, both inpatient and outpatient, from six facilities across the Mount Sinai Health System. Bio*Me* consists of 57,805 selectively-enrolled participants from the Mount Sinai Health System. *All of Us* consists of 413,457 selectively-enrolled participants from across the United States.

### MASLD definition and scoring

For participants who had PDFF measurements available, we defined MASLD cases using PDFF ≥ 5% in addition to at least one of five cardiometabolic criteria.^12^ For participants without PDFF measurements, we defined MASLD cases using ICD-10 codes K75.8 and/or K76.0 in addition to at least one of five cardiometabolic criteria. We defined participants (n = 48) had an ICD-10-based diagnosis but a PDFF < 5% as MASLD cases to account for temporal variations in steatosis.

We established a true “MASLD score” that combines MASLD status with fibrosis severity. As “true” scores, we assigned participants with a PDFF < 5% and no MASLD diagnosis a score of 0, regardless of FIB-4 index. We assigned participants with a PDFF ≥ 5% or a MASLD diagnosis a score from 1 to 3 using their FIB-4 index: 1 for FIB-4 indices < 1.30, 2 for FIB-4 indices between 1.30 and 2.67, and 3 for FIB-4 indices > 2.67. We selected the FIB-4 index due to its superior performance compared to several other non-invasive markers of fibrosis.^13,14^ We then constructed machine learning models using EHR data to predict MASLD scores.

### Participant selection

Across all four biobanks, we employed a consistent filtering strategy (**Figure 1**). We removed patients with missing demographics (gender and age) from all datasets. In the UK Biobank, we selected only participants with clearly defined self-reported ethnicities (i.e., White, Asian, or Black), removing participants of mixed or undefined ethnicities due to ambiguous classification. In the other three datasets, we included all participants regardless of ethnicity and generated MASLD score predictions for participants not of White, Asian, or Black self-reported ethnicity (e.g., Hispanic or Native American) without ethnicity data.

**Figure 1:**
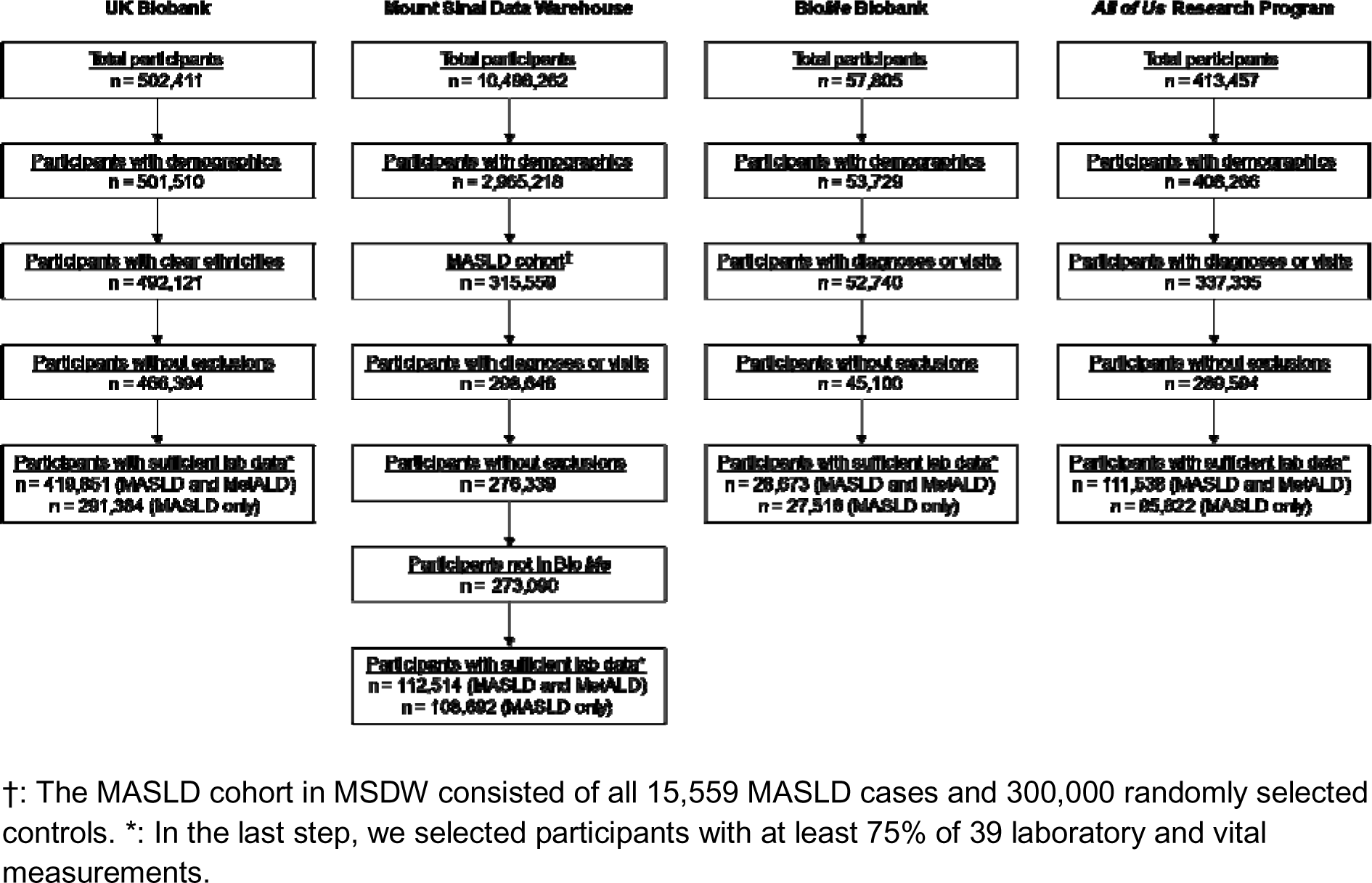
Participant selection procedures across the four biobanks.

To ensure specificity of our model for MASLD, we excluded participants with cryptogenic steatotic liver disease (those who met PDFF or ICD-10 criteria for MASLD but not cardiometabolic criteria) and participants with ICD-10 codes corresponding to a predefined set of non-MASLD liver diseases, alcohol use disorders, and drug use disorders (**Supplemental Table 1**).^15^ To avoid ambiguous cause-effect relationships, we also excluded MASLD cases who had a MASLD-associated outcome (compensated or decompensated cirrhosis, liver transplantation, or liver cancer) prior to the earlier of their MASLD diagnosis or PDFF measurement date (**Supplemental Table 2**).

We performed separate analyses either including or excluding participants meeting alcohol consumption criteria for MetALD. We defined these participants in the UK Biobank as those consuming more than 20 g/day for women and 30 g/day for men; in *All of Us* as those consuming more than two drinks per day; and in MSDW and Bio*Me* as those with daily alcohol consumption.

### Machine learning model

Our model consists of 68 EHR-derived features, including 3 demographic variables, 35 laboratory measurements, 4 vital measurements, 2 social variables, 4 ATC codes, and 20 Elixhauser comorbidities (**Supplemental Table 3-4**). Processing of these features is described in **Supplemental Methods**. We used only EHR data from prior to each participant’s cutoff date, defined as follows: for UK Biobank participants, we used the date of the most recent set of laboratory and vital measurements as the cutoff date, while for Mount Sinai Data Warehouse, Bio*Me*, and All of Us participants, we used the date of the last known diagnosis or patient care encounter.

We constructed the model using LightGBM (version 4.0.0), a gradient boosting framework. Training parameters for all models are listed in **Supplemental Table 5**, and detailed training procedures are available in **Supplemental Methods**. We trained models to minimize mean squared error (MAE) between predicted and actual MASLD scores, and evaluated them in the validation and holdout sets using MAE, root-mean-square error (RMSE), R^2^, and Spearman’s ρ.

To assess model performance on classification tasks (differentiating MASLD cases from controls and differentiating MASLD cases with F3-F4 fibrosis from all other participants), we used area under the receiver-operating curve (AUROC), area under the precision-recall curve (AUPRC), sensitivity, specificity, positive predictive value (PPV), and negative predictive value (NPV). When evaluating AUROC and AUPRC in cohorts without PDFF measurements (MSDW, Bio*Me*, All of Us), we addressed the issue of severe MASLD underdiagnosis based on ICD-10 codes by creating balanced datasets with MASLD score distributions matching those of the training dataset.

### Statistical analyses

We performed all statistical analyses in Python 3.11.4 and set the significance level to 0.05 for all tests. To evaluate clinical associations, we segmented participants’ predicted scores into quintiles separately for each biobank; we then used either statsmodels (version 0.13.5) to calculate odds ratios or lifelines (version 0.27.4) to perform Cox proportional hazards regression between the quintiles and each comorbidity or outcome. We also performed Cox regression to evaluate the association between quintiles and mortality in UK Biobank and Bio*Me*; both MSDW and *All of Us*. In all analyses, we adjusted for age, gender, and ethnicity. Importantly, to prevent data leakage, we excluded all outcomes from the diagnoses used as inputs for machine learning models.

## Results

### Participants

We trained machine learning models using EHRs from 37,212 participants in the UK Biobank: 33,148 with PDFF measurements, and 4,064 without PDFF but with a MASLD diagnosis. Of these participants, 19,538 [52.55%] were female; 36,338 [97.7%] were White, 590 [1.6%] were Asian, and 284 [0.8%] were Black; the median age was 57.2 [IQR 12.3]; and 13,055 [35%] were MASLD cases (**Supplemental Table 6**). For the 33,148 participants with PDFF measurements, the median was 3.0% [IQR 3.1%].

Subsequently, we used models to make predictions in four different cohorts with 635,364 total participants, including 252,725 participants from across the United States **(Supplemental Table 7)**. The first cohort included 382,639 UK Biobank participants not included in the training set. Of these, 210,738 [55.1%] were female; 367,427 [96.0%] were White, 8,974 [2.3%] were Asian, and 6,238 [1.6%] were Black; and the median age was 58.8 [IQR 13.2]. The second cohort included 112,514 participants from the Mount Sinai Data Warehouse. Of these participants, 64,730 [57.5%] were female; 77,184 [68.6%] were White, 23,414 [20.8%] were Black, and 11,202 [10.0%] were Asian; and the median age was 55.6 [IQR 34.4]. The third cohort included 28,673 participants from Bio*Me*. Of these participants, 17,780 [62.0%] were female; 11,263 [39.3%] were White, 6,455 [22.5%] were Black, and 1,056 [3.7%] were Asian; and the median age was 59.3 [IQR 27.3]. The fourth cohort included 111,538 participants from *All of Us*. Of these participants, 72,690 [65.2%] were female; 68,200 [61.1%] were White, 15,987 [14.3%] were Black, and 3,269 [2.9%] were Asian; and the median age was 59.1 [IQR 27.8].

### Performance metrics

Our machine learning models are gradient boosting regression models that assign participants a predicted MASLD score from 0 to 3. 0 suggests no MASLD, 1 suggests MASLD with F0-F2 fibrosis, 2 suggests MASLD with indeterminate fibrosis, and 3 suggests MASLD with F3-F4 fibrosis. However, we explicitly aim for this score to be continuous, with a higher score suggesting MASLD with more severe fibrosis.

Recognizing the overlap between and changing definitions of MASLD and MetALD, we trained separate models and performed separate analyses where participants meeting alcohol consumption criteria for MetALD were either included or excluded. Comparing predicted and actual MASLD scores in the holdout set, models showed minimal error rates with RMSEs of 0.61 (0.61-0.61) and 0.59 (0.59-0.59), as well as MAEs of 0.45 (0.45-0.45) and 0.43 (0.43-0.43) when MetALD participants were included or excluded, respectively (**Supplemental Table 8**). We observed that the distribution of predicted MASLD scores in external test sets closely mirrored those in the holdout set (**Figure 2a-e**). However, many participants assigned a MASLD score of 0 were predicted to have scores above 0.5 (192,253 [50.2%] in the UK Biobank prediction set; 56,299 [50.5%] in MSDW; 17,126 [62.9%] in Bio*Me*; and 70,742 [68.6%] in *All of Us*, which may be attributed to MASLD underdiagnosis.

**Figure 2:**
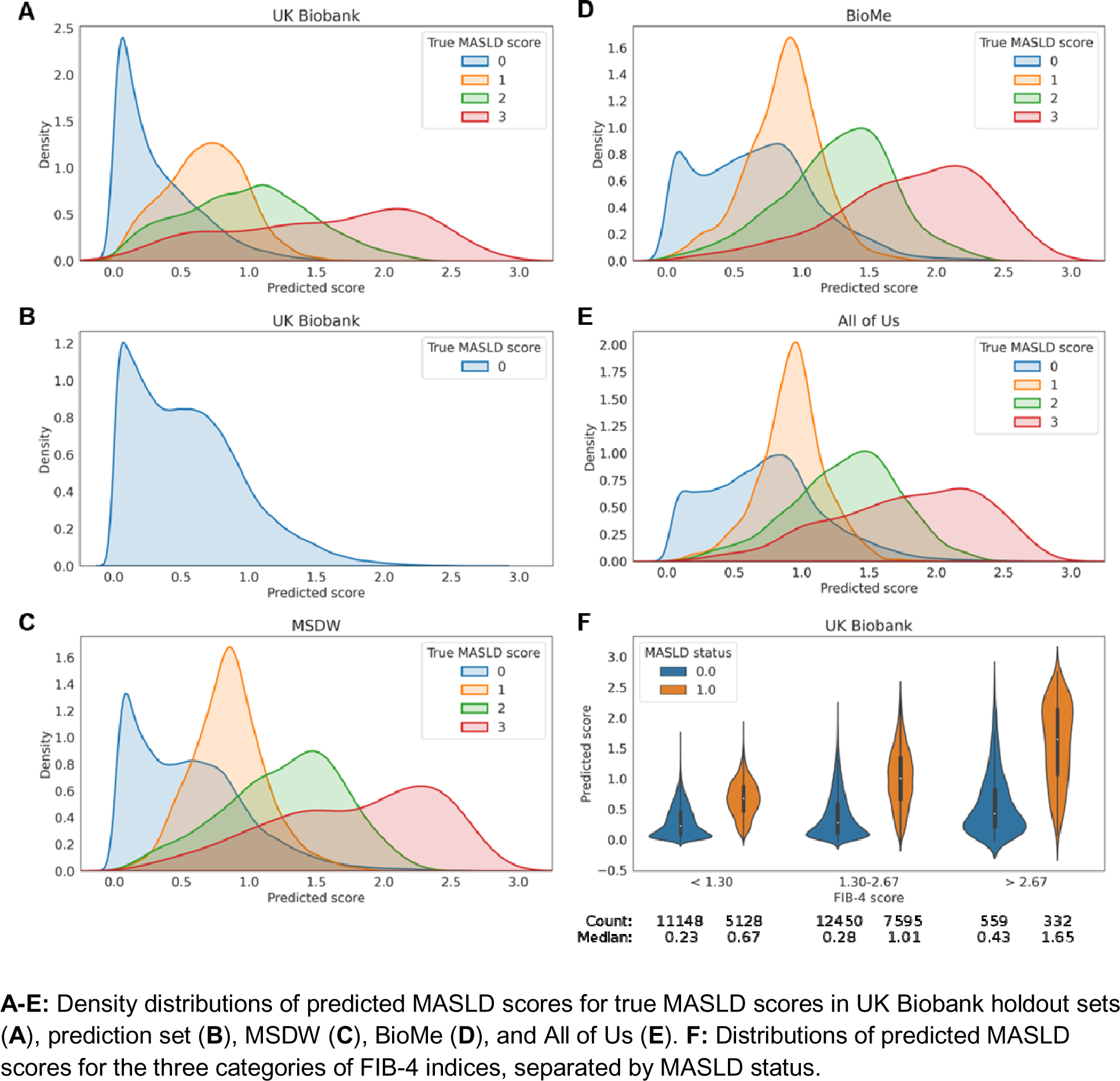
Distribution of predicted scores in holdout and external validation cohorts.

We evaluated model performance on two classification tasks: (1) differentiating individuals with and without MASLD (i.e., score ≥ 1 from score < 1; hereafter, MASLD diagnosis), and (2) differentiating individuals with both MASLD and F3-F4 fibrosis from all other individuals (i.e., score = 3 from score < 3; hereafter, fibrosis identification). In the holdout set, our models achieved an AUROC of 0.83 (0.83-0.83) for MASLD diagnosis and an AUROC of 0.91 (0.90-0.92) for fibrosis identification (**Table 1**). They outperformed the hepatic steatosis index (HSI), which achieved an AUROC of 0.80 (0.80-0.80) for MASLD diagnosis and an AUROC of 0.61 (0.60-0.62) for fibrosis identification in the same holdout set (**Supplemental Table 9**). Notably, in all three FIB-4 classes (FIB-4 < 1.30, 1.30 - 2.67, and > 2.67), predicted MASLD scores were significantly higher (p < 0.001) for participants with MASLD compared to those without (**Figure 2f**), indicating the model distinguishes MASLD-associated fibrosis from fibrosis due to other causes. This is useful as the FIB-4 index alone does not discriminate between MASLD cases and controls, with an AUROC of 0.45 (0.45-0.45) and AUPRC of 0.33 (0.33-0.33) for MASLD diagnosis in the holdout set (**Supplemental Table 10**).

**Table 1:** Classification metrics for models evaluated on holdout and external validation sets.

| Task | Dataset | AUROC | AUPRC | Proportion |
| --- | --- | --- | --- | --- |
| Holdout (MASLD and MetALD) |  |  |  |  |
| MASLD diagnosis | UK Biobank | 0.83 (0.83-0.83) | 0.71 (0.71-0.71) | 0.35 |
| Fibrosis identification |  | 0.91 (0.90-0.92) | 0.36 (0.34-0.38) | 0.01 |
| Predictions (MASLD and MetALD) |  |  |  |  |
| MASLD diagnosis | MSDW | 0.79 (0.78-0.80) | 0.62 (0.61-0.63) | 0.35 |
|  | BioMe | 0.74 (0.73-0.75) | 0.56 (0.55-0.57) |  |
|  | All of Us | 0.74 (0.74-0.74) | 0.55 (0.55-0.55) |  |
| Fibrosis identification | MSDW | 0.89 (0.87-0.91) | 0.37 (0.29-0.45) | 0.01 |
|  | BioMe | 0.92 (0.91-0.93) | 0.30 (0.24-0.36) |  |
|  | All of Us | 0.89 (0.89-0.90) | 0.24 (0.22-0.26) |  |
| Holdout (MASLD only) |  |  |  |  |
| MASLD diagnosis | UK Biobank | 0.84 (0.84-0.84) | 0.71 (0.71-0.71) | 0.34 |
| Fibrosis identification |  | 0.90 (0.89-0.91) | 0.40 (0.38-0.42) | 0.01 |
| Predictions (MASLD only) |  |  |  |  |
| MASLD diagnosis | MSDW | 0.80 (0.80-0.80) | 0.61 (0.60-0.62) | 0.34 |
|  | BioMe | 0.73 (0.72-0.74) | 0.53 (0.52-0.54) |  |
|  | All of Us | 0.74 (0.74-0.75) | 0.54 (0.54-0.54) |  |
| Fibrosis identification | MSDW | 0.91 (0.89-0.93) | 0.38 (0.31-0.45) | 0.01 |
|  | BioMe | 0.90 (0.88-0.92) | 0.24 (0.19-0.29) |  |
|  | All of Us | 0.89 (0.88-0.90) | 0.26 (0.24-0.27) |  |

**Interpretation:** AUROC evaluates the trade-off between sensitivity (ability to identify true positives) and specificity (ability to avoid false positives) across multiple thresholds, with 1.0 denoting perfect discrimination and 0.5 denoting random performance. AUPRC evaluates a model's effectiveness at predicting positives over a range of thresholds by analyzing the balance between precision (correctly predicted positives out of all predicted positives) and recall (correctly predicted positives out of all actual positives). **Abbreviations:** AUROC: area under the precision recall curve. AUPRC: area under the precision recall curve.

We also evaluated task performance at different thresholds. A MASLD score threshold of 0.50 balances sensitivity and specificity for MASLD diagnosis, yielding 0.77 sensitivity, 0.74 specificity, 0.61 PPV, and 0.85 NPV (**Supplemental Table 11**), whereas FIB-4 fails to accurately identify MASLD cases at either its 1.30 or 2.67 thresholds. MASLD score thresholds between 0.75 and 1.25 offer balanced performance on both tasks and may be ideal for a clinical alert system, where patients with scores above the threshold are deemed high-risk and are recommended for referral and further workup. For fibrosis identification, at equivalent thresholds of 2.00 (MASLD score) and 2.67 (FIB-4 index), FIB-4 has higher sensitivity than MASLD score (1.00 versus 0.32), which is expected as we define fibrosis using the FIB-4 index in this study (**Supplemental Table 12**). However, FIB-4 has significantly lower PPV (0.37 versus 0.48) due to increased false positives (i.e., identifying F3-F4 fibrosis not due to MASLD); this is consistent with our model, but not FIB-4, being able to identify MASLD-associated fibrosis.

Next, we analyzed model performance on these two classification tasks in participants without PDFF available. Because of the underdiagnosis of MASLD among these participants when relying on ICD-10 codes, we created balanced cohorts for each biobank that matched the MASLD score distributions of the training set. Across all four biobanks, models achieved consistent performance in both tasks, with AUROCs ranging from 0.73-0.80 for MASLD diagnosis and 0.89-0.92 for fibrosis classification (**Table 1**), despite there being large numbers of undiagnosed MASLD cases in each of these datasets.

Because the UK Biobank has limited diversity, with participants being majority White and aged 40-69, we also examined the ability of our models to extrapolate to diverse populations by performing a subgroup analysis. In MSDW and *All of Us*, performance was generally consistent across different genders, ethnicities, and age groups (**Supplemental Tables 13-15**). However, there was reduced performance for certain subsets with limited representation in the training set, including Black participants (0.01-0.05 decrease in AUROC in MSDW and *All of Us*) and participants over 69 years of age (0.05-0.13 decrease in AUROC in MSDW and *All of Us*).

### Feature importance

SHAP analysis demonstrated that the 10 most important features for our model were well-known risk factors for MASLD (**Extended Data Figure 1**). BMI and triglycerides were the first and second most important features, while the four variables used in the FIB-4 index (platelet count, ALT, AST, and age) were the third to sixth most important features, respectively. Diagnostic history was also important, with hypertension, diabetes, obesity, and chronic pulmonary disease being the 6th, 9th, 11th, and 16th most important features, respectively. However, our model also identified several features not previously associated with MASLD, such as total protein, albumin, and monocyte count, all of which were positively correlated with SHAP values.

### Clinical associations

Predicted MASLD scores were significantly associated with MASLD outcomes and MASLD comorbidities in all four biobanks, both when including and excluding MetALD participants. MASLD scores had significant associations with several major MASLD comorbidities, including ischemic heart disease, atrial fibrillation, heart failure, chronic kidney disease, type 2 diabetes, and obstructive sleep apnea (**Table 2**); associations with the latter four comorbidities were also present separately among cases and controls (**Supplemental Tables 16-17**). We observed the highest odds ratios (ORs) for type 2 diabetes, with ORs per quintile increase in predicted score ranging from 2.39 (2.35-2.44) in MSDW to 3.93 (3.87-4.00) in the UK Biobank. To determine the specificity of our model, we tested associations between predicted score and MASLD diagnosis based on ICD-10 coding. Here, each quintile increase in model score was associated with ORs ranging from 2.24 (2.19-2.26) in All of Us to 3.19 (2.97-3.44) in MSDW.

**Table 2:**
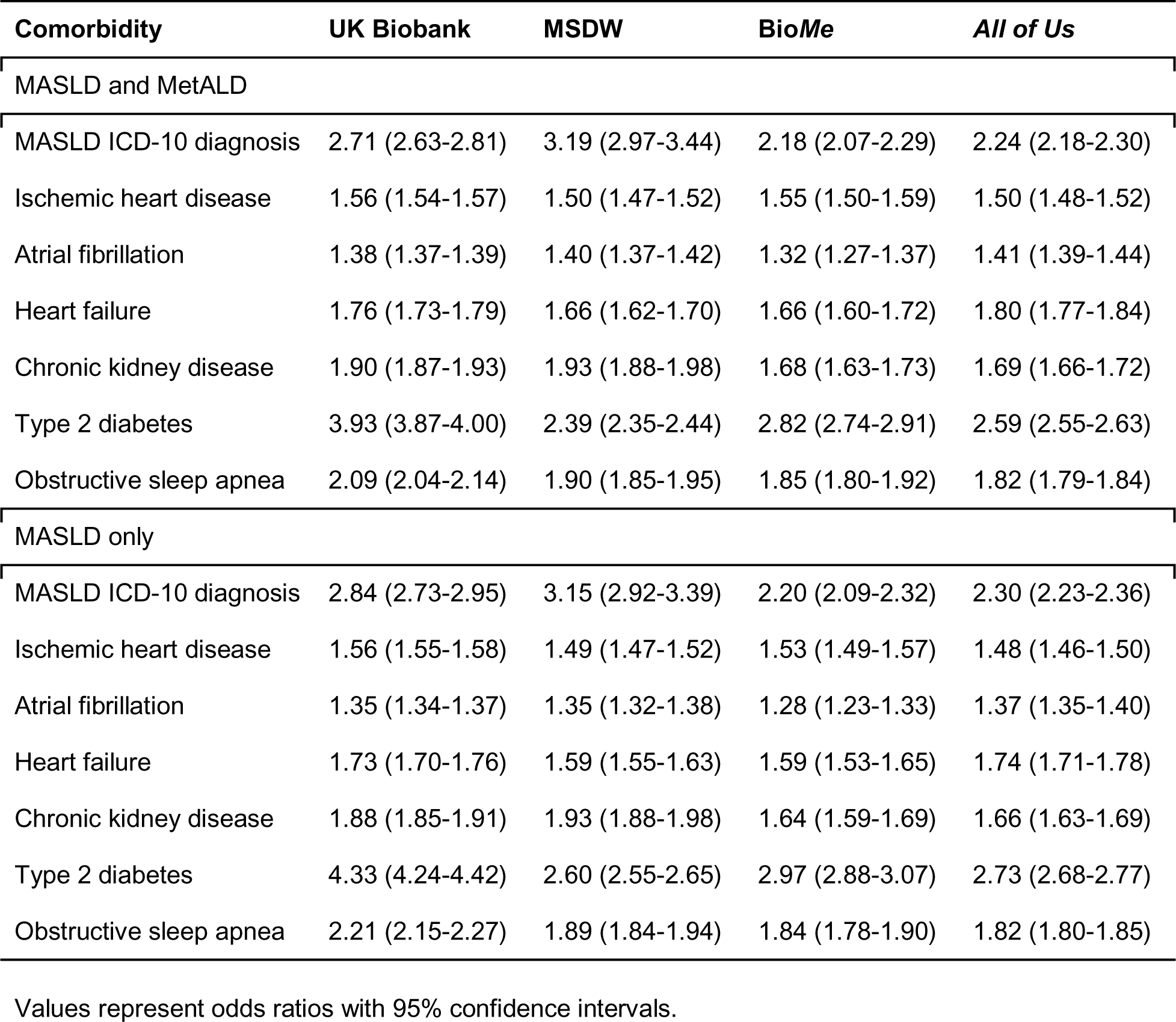
Odds ratios per quintile increase in predicted score for MASLD comorbidities among all participants.

| **Comorbidity** | **UK Biobank** | **MSDW** | **BioMe** | **All of Us** |
| --- | --- | --- | --- | --- |
| MASLD and MetALD |  |  |  |  |
| MASLD ICD-10 diagnosis | 2.71 (2.63-2.81) | 3.19 (2.97-3.44) | 2.18 (2.07-2.29) | 2.24 (2.18-2.30) |
| Ischemic heart disease | 1.56 (1.54-1.57) | 1.50 (1.47-1.52) | 1.55 (1.50-1.59) | 1.50 (1.48-1.52) |
| Atrial fibrillation | 1.38 (1.37-1.39) | 1.40 (1.37-1.42) | 1.32 (1.27-1.37) | 1.41 (1.39-1.44) |
| Heart failure | 1.76 (1.73-1.79) | 1.66 (1.62-1.70) | 1.66 (1.60-1.72) | 1.80 (1.77-1.84) |
| Chronic kidney disease | 1.90 (1.87-1.93) | 1.93 (1.88-1.98) | 1.68 (1.63-1.73) | 1.69 (1.66-1.72) |
| Type 2 diabetes | 3.93 (3.87-4.00) | 2.39 (2.35-2.44) | 2.82 (2.74-2.91) | 2.59 (2.55-2.63) |
| Obstructive sleep apnea | 2.09 (2.04-2.14) | 1.90 (1.85-1.95) | 1.85 (1.80-1.92) | 1.82 (1.79-1.84) |
| MASLD only |  |  |  |  |
| MASLD ICD-10 diagnosis | 2.84 (2.73-2.95) | 3.15 (2.92-3.39) | 2.20 (2.09-2.32) | 2.30 (2.23-2.36) |
| Ischemic heart disease | 1.56 (1.55-1.58) | 1.49 (1.47-1.52) | 1.53 (1.49-1.57) | 1.48 (1.46-1.50) |
| Atrial fibrillation | 1.35 (1.34-1.37) | 1.35 (1.32-1.38) | 1.28 (1.23-1.33) | 1.37 (1.35-1.40) |
| Heart failure | 1.73 (1.70-1.76) | 1.59 (1.55-1.63) | 1.59 (1.53-1.65) | 1.74 (1.71-1.78) |
| Chronic kidney disease | 1.88 (1.85-1.91) | 1.93 (1.88-1.98) | 1.64 (1.59-1.69) | 1.66 (1.63-1.69) |
| Type 2 diabetes | 4.33 (4.24-4.42) | 2.60 (2.55-2.65) | 2.97 (2.88-3.07) | 2.73 (2.68-2.77) |
| Obstructive sleep apnea | 2.21 (2.15-2.27) | 1.89 (1.84-1.94) | 1.84 (1.78-1.90) | 1.82 (1.80-1.85) |

Values represent odds ratios with 95% confidence intervals.

We also examined the major outcomes of MASLD, including compensated and decompensated cirrhosis, liver cancer, and liver transplantation. Among all UK Biobank participants, over a 15-year follow-up period, each quintile increase in predicted MASLD score was associated with HRs of 1.10 (1.08-1.12), 1.11 (1.09-1.13), and 1.15 (1.13-1.17) for a new diagnosis of compensated, decompensated, and any cirrhosis, respectively (**Table 3**); these associations were present separately among MASLD cases and controls. Among only MASLD cases, HRs for these outcomes increased to 1.54 (1.36-1.73), 1.37 (1.21-1.54), and 1.53 (1.37-1.71), respectively. Similarly, among a subset of 48,619 MSDW participants with complete data from 3-6 years prior to the cutoff date, we observed that over a 3-year follow-up period, each quintile increase was associated with HRs of 1.10 (1.03-1.16), 1.08 (1.02-1.14), and 1.11 (1.05-1.17) for new diagnoses of compensated, decompensated, and any cirrhosis, respectively (**Supplemental Table 18**). Likewise, these associations were present among both cases and controls, and among cases, HRs for these outcomes increased to 1.87 (1.13-3.08), 1.98 (1.10-3.56), and 1.91 (1.15-3.15), respectively.

**Table 3:** Hazard ratios per quintile increase in predicted score for MASLD outcomes.

|  | Cases and controls |  | Only cases |  | Only controls |  |
| --- | --- | --- | --- | --- | --- | --- |
| Outcome | Count | HR (95% CI) | Count | HR (95% CI) | Count | HR (95% CI) |
| MASLD and MetALD |  |  |  |  |  |  |
| Compensated cirrhosis | 370352 | 1.10 (1.08-1.12) | 11825 | 1.54 (1.36-1.73) | 358527 | 1.07 (1.05-1.10) |
| Decompensated cirrhosis | 370178 | 1.11 (1.09-1.13) | 11827 | 1.37 (1.21-1.54) | 358351 | 1.10 (1.08-1.13) |
| Any cirrhosis | 370120 | 1.15 (1.13-1.17) | 11822 | 1.53 (1.37-1.71) | 358298 | 1.13 (1.11-1.15) |
| Liver cancer | 370414 | 1.03 (1.00-1.05) | 11823 | 1.34 (1.18-1.53) | 358591 | 1.01 (0.99-1.03) |
| Liver transplant | 370419 | 1.03 (1.01-1.06) | 11832 | 1.16 (1.01-1.34) | 358587 | 1.03 (1.01-1.05) |
| MASLD only |  |  |  |  |  |  |
| Compensated cirrhosis | 257266 | 1.10 (1.07-1.13) | 8165 | 1.59 (1.38-1.83) | 249101 | 1.07 (1.04-1.10) |
| Decompensated cirrhosis | 257135 | 1.11 (1.09-1.14) | 8166 | 1.40 (1.21-1.62) | 248969 | 1.10 (1.08-1.13) |
| Any cirrhosis | 257090 | 1.15 (1.13-1.17) | 8162 | 1.57 (1.38-1.80) | 248928 | 1.13 (1.10-1.15) |
| Liver cancer | 257316 | 1.03 (1.00-1.06) | 8165 | 1.35 (1.14-1.58) | 249151 | 1.01 (0.98-1.04) |
| Liver transplant | 257317 | 1.03 (1.00-1.06) | 8171 | 1.15 (0.96-1.36) | 249146 | 1.03 (1.00-1.05) |

“Count” represents the number of participants in each analysis. For each outcome, participants with an outcome prior to their cutoff date were excluded. Values represent hazard ratios with 95% confidence intervals. **Abbreviations:** HR: hazard ratio. CI: confidence interval.

In Bio*Me* and *All of Us*, we calculated ORs rather than HRs due to limited follow-up periods, comparing them to ORs in UK Biobank and MSDW. We examined whether a higher predicted MASLD score was associated with increased likelihood of a past diagnosis of a MASLD outcome, and observed that they were indeed significantly associated with all outcomes: ORs per quintile increase in predicted score ranged from 1.35 (1.30-1.39) in *All of Us* to 1.90 (1.79-2.03) in MSDW for any cirrhosis; 1.57 (1.35-1.81) in *All of Us* to 2.11 (1.86-2.39) in MSDW for liver cancer; and 1.65 (1.35-2.03) in *All of Us* to 2.14 (1.56-2.93) in UK Biobank for liver transplant (**Supplemental Table 19**). Associations were also present separately among cases and controls, with higher ORs among cases for all outcomes (**Supplemental Tables 20-21**).

### Mortality associations

MASLD patients are known to have significantly higher all-cause mortality, with an HR of 1.34 (1.17-1.54) across fourteen studies.^16^ Consistent with this, predicted MASLD scores were also significantly associated with all-cause mortality in the UK Biobank and Bio*Me*, as well as disease-specific mortality in the UK Biobank. Examining all-cause mortality in the UK Biobank, each quintile increase in predicted score resulted in HRs of 1.26 (1.25-1.27) among all participants, 1.24 (1.10-1.39) among cases, and 1.26 (1.25-1.27) among controls (**Figure 3a-b**; **Supplemental Figure 1a-b**). Furthermore, examining disease-specific mortality, each quintile increase resulted in HRs of 1.33 (1.31-1.35) for mortality due to cardiovascular disease, 1.23 (1.20-1.25) for mortality due to diabetes, and 1.22 (1.21-1.24) for mortality due to extrahepatic cancer (**Supplemental Table 22**), all of which are known common causes of death in MASLD patients.^17^ Significant associations for all-cause mortality also present in Bio*Me*, where each quintile increase in predicted score resulted in HRs of 1.13 (1.08-1.19) among all participants and 1.13 (1.07-1.19) among controls (**Figure 3c-d**; **Supplemental Figure 1c-d**).

**Figure 3:**
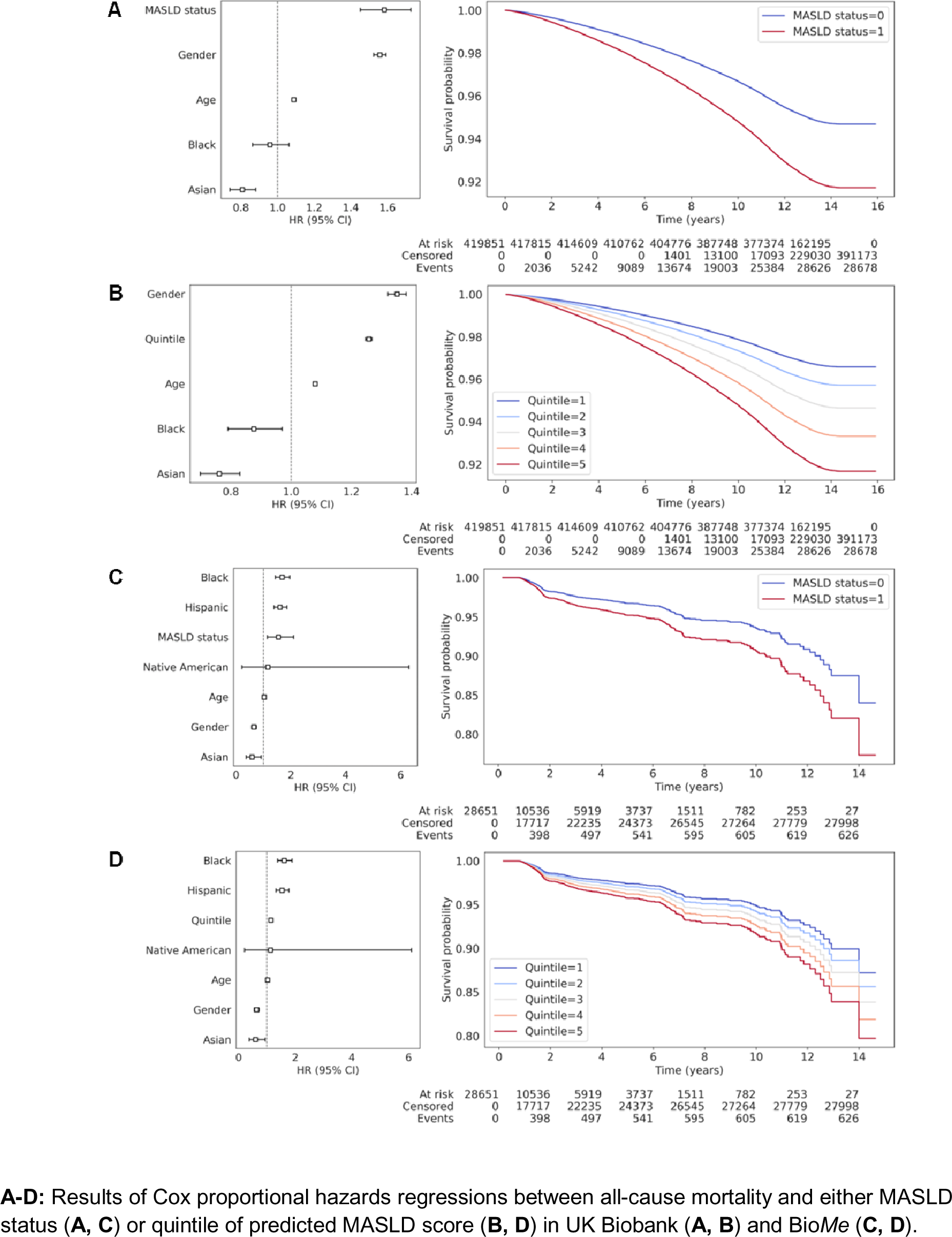
Association of predicted MASLD scores with mortality risk in UK Biobank and Bio*Me*.

### Biomarker associations

Several laboratory biomarkers not included in our model have been independently associated with MASLD. For example, increased GGT and decreased CCR are markers and/or predictors of advanced fibrosis in MASLD;^18,19^ inflammatory markers like ESR and CRP may reflect the chronic low-grade inflammation in MASLD patients;^20^ while estradiol which may be protective against MASLD.^21,22^ We thus performed linear regression analyses between predicted scores and these biomarkers in UK Biobank, MSDW, and *All of Us*, and observed consistent results across biobanks and between cases and controls (**Supplemental Tables 23-25**). For inflammatory markers, predicted scores were positively correlated with C reactive protein (CRP), with βs of 1.82 in UK Biobank, 2.42 in MSDW, and 1.40 in *All of Us*; as well as erythrocyte sedimentation rate (ESR), with βs of 18.75 in MSDW and 10.88 in *All of Us*. Other biomarkers included gamma glutamyltransferase (GGT), with βs of 32.25 in UK Biobank, 80.55 in MSDW, and 61.77 in *All of Us*; creatinine/cystatin C ratio (CCR), with a β of -62.92 in UK Biobank; and estradiol, with a β of -223.52 in UK Biobank. All correlations were significant to p < 0.001.

### Underdiagnosis identification

We observed widespread underdiagnosis of MASLD based on administrative coding. Among the 33,148 participants with PDFF measurements, 27.1% had either a MASLD diagnosis or a PDFF > 5%. In contrast, among the other four datasets, the percent of participants with a MASLD diagnosis ranged from 7.5% in *All of Us* to 1.0% in MSDW (**Supplemental Table 7**), significantly below prevalence estimates of 25-40% in the United States.^1,7,23^

To assess our model’s efficacy in detecting MASLD underdiagnosis, a hepatologist specialized in MASLD reviewed charts of 36 Bio*Me* participants without a MASLD diagnosis (**Supplemental Table 26**). We organized participants into 12 sets of three, matching participants in each set by age, gender, and ethnicity. Within each set, we selected participants with predicted MASLD scores closest to 0, 1, or 2. Of the 12 participants with scores ≈ 0, 11 had no evidence of MASLD, and one had possible MASLD without fibrosis. For those with scores ≈ 1, one had definitive MASLD based on ultrasound, and two had possible MASLD. Among participants with scores ≈ 2, one had MASLD with F0-F1 fibrosis based on elastography, one had MASLD with cirrhosis based on MRI, three had possible MASLD with fibrosis, and two had possible MASLD with undetermined fibrosis. Further, ordinal logistic regression indicated that predicted MASLD score was significantly associated with an increased confidence of hepatologist-identified MASLD, with an OR per unit increase in score of 4.99 (1.60-15.55) (**Supplemental Table 27**).

## Discussion

In the pursuit of precision medicine, the application of computational models to large-scale EHR data can revolutionize our understanding of complex diseases. MASLD is one such disease: it has a wide spectrum of severity, from being asymptomatic to causing liver failure, and is severely underdiagnosed, which hampers optimal management strategies.^24^ Underdiagnosis of MASLD reflects broader issues in medical practice where conditions elude diagnosis due to difficulty of accessing diagnostic tools or a general lack of awareness. Addressing these issues, we described here a machine learning model trained and evaluated on four diverse biobanks to simultaneously assess both MASLD presence and severity.

Our model was well-calibrated and performed well across four replication cohorts with varying health systems, ethnicities, population and disease ascertainment, and country of origin. This suggests robustness to these differences and versatility across different settings. Importantly, our model distinguished MASLD-associated fibrosis from fibrosis due to other causes: in all three FIB-4 index bins, MASLD cases had significantly higher predicted scores than controls. Although performance was slightly reduced in the replication cohorts compared to the holdout set, this could be due to the significant underdiagnosis of MASLD among these cohorts, resulting in inaccurate case/control definitions. Indeed, expert chart review demonstrated that our model could identify individuals without a recorded diagnosis of MASLD but had imaging-confirmed MASLD with fibrosis or cirrhosis.

To enable population-wide screening, we designed our model to be portable across healthcare settings and use existing data. As such, it uses only commonly measured laboratory measurements (those part of a complete metabolic panel, complete blood count, and a lipid panel) and a limited number of Elixhauser comorbidities that can be readily extracted from patients’ diagnostic histories. This contrasts scores such as Hepamet, Fibrometer, and FibroMax, which require specialized labs (e.g., gamma glutamyltransferase and alpha-2-macroglobulin) that patients are unlikely to already have available. Our model can also be applied to patients with missing measurements because, just as we imputed missing data separately for each cohort, health systems can perform imputation using their own patient data.

Further, given that 30% of the population has MASLD, it is important that clinicians can use our model to prioritize evaluation and treatment for patients with severe disease and/or who are most at risk for progression, with the goal of minimizing costs, labor, and unnecessary use of diagnostic equipment. To validate this ability of our models, we demonstrated that in both UK Biobank and MSDW, scores were significantly associated with higher hazards for newly developing both compensated cirrhosis and decompensated cirrhosis in a gradated manner; and that in both UK Biobank and Bio*Me*, scores were significantly associated with increased risk of all-cause mortality. Our model thus contrasts both existing steatosis scores like the HSI, which fails to capture fibrosis severity, and fibrosis scores like FIB-4, which identifies fibrosis but does not distinguish MASLD-associated fibrosis from other etiologies.

As additional evidence of clinical validity, we found significant associations between predicted MASLD scores and prior diagnoses of several MASLD comorbidities associated with progression to MASH and fibrosis.^25^ In all four biobanks, increased scores were also significantly associated with prior diagnoses of cirrhosis and liver cancer, suggesting our model may be useful in identifying MASLD patients who have already progressed to advanced disease but have not yet been diagnosed. Surprisingly, there was also an association with prior liver transplant; this may reflect the persistence/recurrence of MASLD and metabolic dysfunction after transplantation.^26^

Despite these strengths, our study has several limitations. First, we trained the model using a predominantly white European cohort, and while subset analyses suggested that the model generalizes well to diverse populations, there are nonetheless slight decreases in performance among demographics underrepresented in the training data, emphasizing the need for more diverse datasets. Second, for some MASLD patients who progress to cirrhosis, their liver fat percentage decreases below 5%; while rare, these patients may be underrepresented in our model, as we partially defined MASLD as PDFF ≥ 5%. Third, without elastography data, we used the FIB-4 index as a proxy for liver fibrosis among MASLD patients. The FIB-4 index is an imperfect but nevertheless excellent predictor of advanced fibrosis among MASLD patients: at the lower cutoff of 1.30 used in this study, the FIB-4 index has a negative predictive value of 0.90 for ruling out advanced fibrosis, while at the higher cutoff of 2.67, it has a positive predictive value of 0.80 for ruling in advanced fibrosis.^13,14^ We also note that the goal of our model is not to replace diagnostic tools (e.g., elastography, imaging, and biopsy), but rather to provide a more useful MASLD screening tool among the general population that increases access to and efficient usage of these tools.

## Data sharing statement

Further information about the MSDW and Bio*Me* datasets is available at https://labs.icahn.mssm.edu/msdw/ and https://icahn.mssm.edu/research/ipm/programs/biome-biobank/, respectively. The UK Biobank dataset can be accessed by applying through the Access Management System at https://bbams.ndph.ox.ac.uk/ams/. The *All of Us* dataset can be accessed by applying through the *All of Us* Research Hub at https://www.researchallofus.org/. Code used for the analyses have been deposited at https://doi.org/10.17632/78gc6dpwvm.1.

## Data Availability

Further information about the MSDW and BioMe datasets is available at https://labs.icahn.mssm.edu/msdw/ and https://icahn.mssm.edu/research/ipm/programs/biome-biobank/, respectively. The UK Biobank dataset can be accessed by applying through the Access Management System at https://bbams.ndph.ox.ac.uk/ams/. The All of Us dataset can be accessed by applying through the All of Us Research Hub at https://www.researchallofus.org/. Code used for the analyses have been deposited at https://doi.org/10.17632/78gc6dpwvm.1.

## Acknowledgements

This work was supported in part through the Mount Sinai Data Warehouse (MSDW) resources and staff expertise provided by Scientific Computing and Data at the Icahn School of Medicine at Mount Sinai. This work required the use of the *All of Us* Research Program, which is supported by the Office of the Director of the National Institutes of Health (NIH) and would not be possible without the partnership of its participants.

## Extended Data

**Extended Data Figure 1:**
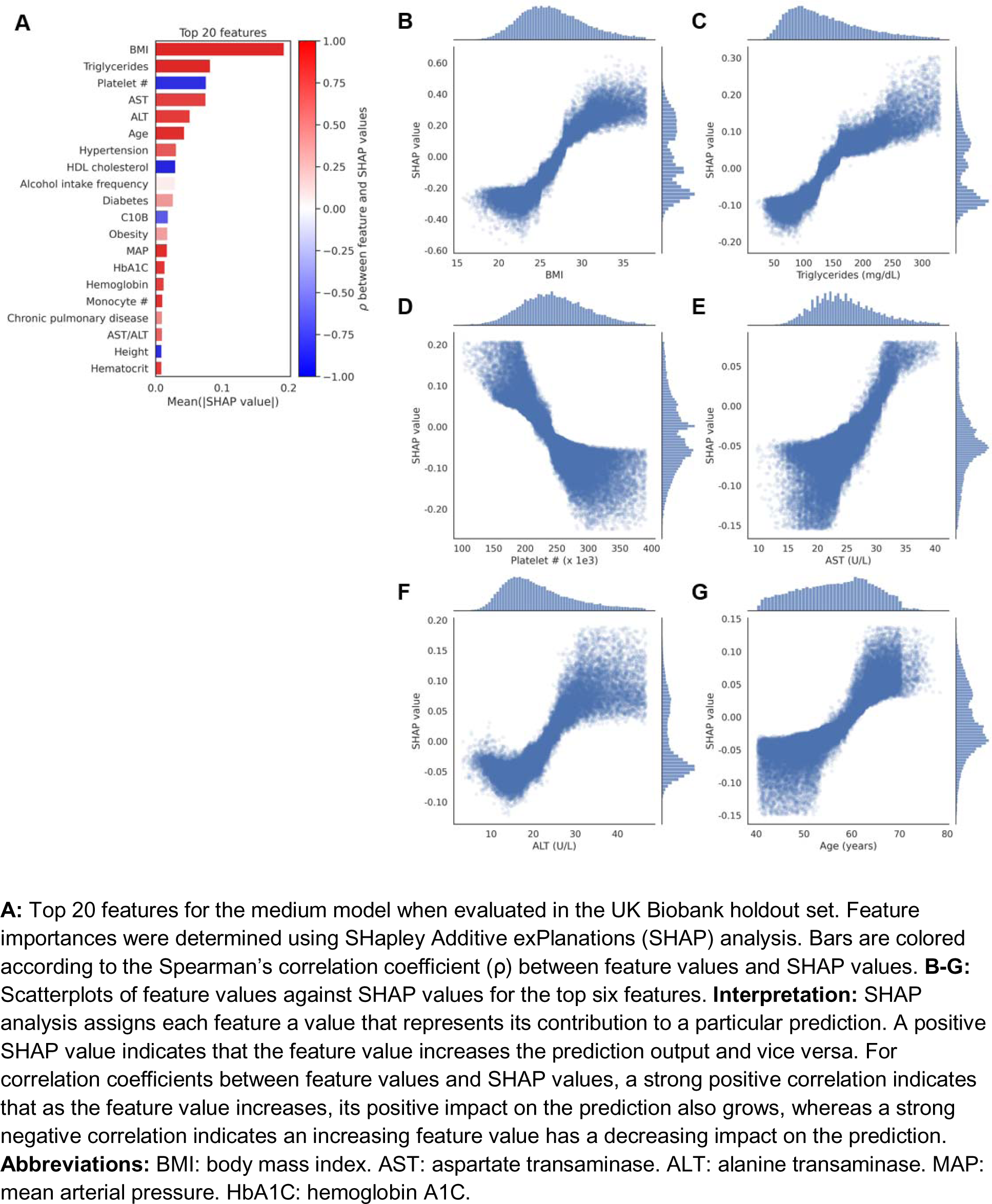
Analysis of important features for machine learning models.

