## Supplemental Figures and Tables for "Machine Learning Enables Single-Score Assessment of MASLD Presence and Severity"

### Supplemental Methods

**Feature processing**

For each type of laboratory and vital measurement, we used the median of all measurements from up to five years prior to the cutoff date. We selected only patients with alanine aminotransferase (ALT), aspartate aminotransferase (AST), body mass index (BMI), and platelet counts available, as these were required for the FIB-4 score, as well as 75% of the 39 frequently measured laboratory and vital measurements (**Supplemental Table 3**). We imputed remaining missing features using the IterativeImputer function of scikit-learn (version 1.3.0), stopping upon reaching the default tolerance of 1e-3. In each iteration of imputation, this function uses Bayesian ridge regression to model each missing feature as a function of other features in a round-robin fashion, moving from features with fewest missing values to most. We performed imputation separately for each of the four cohorts to prevent data leakage.

To incorporate diagnostic history into our model, we removed all ICD-10 codes associated with MASLD or MASLD outcomes (**Supplemental Table 2**) and converted the remaining ICD-10 codes into 20 Elixhauser comorbidities (**Supplemental Table 4**). Each comorbidity was encoded as either 0 (not present) or 1 (present).

For medication usage, we converted all medication data in each biobank to ATC codes. Because the UK Biobank does not specify medication usage dates, we used binary encoding, with 1 indicating usage of medications within an ATC code at any point prior to the cutoff date. We used four ATC codes as features: A02B (drugs for peptic ulcer and gastro-esophageal reflux disease), A10B (blood glucose lowering drugs, excluding insulins), C10A (lipid modifying agents, plain), and C10B (lipid modifying agents, combinations). These were selected based on both biological plausibility and feature importance.

We encoded alcohol intake frequency on an ordinal 0 to 4 scale. 0 indicates no alcohol intake; 1 indicates less than monthly alcohol intake; 2 indicates monthly alcohol intake; 3 indicates weekly alcohol intake; and 4 indicates daily alcohol intake. We encoded smoking status as a binary feature, with 0 indicating no and 1 indicating yes.

**Model training procedure**

To train our models, we used a cohort of UK Biobank participants with either PDFF measurements (n = 33,148) or a MASLD diagnosis based on ICD-10 coding (n = 4,064). We included the latter group as ICD-10 codes have high positive predictive values for MASLD.^1^ We partitioned this cohort into ten equally sized subsets, or folds. We used a nested cross-validation approach to ensure robust model training and evaluation. In this process, each fold served as a holdout set once, while the other nine folds were further divided into ten smaller folds for training and validation. Specifically, within these nine folds, one fold was used for validation and the remaining nine for training. After model training using its designated validation set, each model was tested on the holdout set, and then used to make predictions on all participants not included in the training cohort. This process was iterated ten times for each initial fold, resulting in a total of 100 iterations (10 outer folds x 10 inner folds). These iterations were used to generate confidence intervals for all metrics.

**Chart review**

To determine if our model could identify underdiagnosis, a hepatologist with MASLD expertise (M.B.) performed retrospective chart review in 12 sets of three matched Bio*Me* participants (36 participants total). The Icahn School of Medicine Institutional Review Board provided approval under study 10-00158. To generate each set of matches, we first randomly selected a participant from Bio*Me* and extracted their age, gender, and ethnicity. We then identified participants with the same gender and ethnicity and an age difference of less than one year. From this group, we selected the three participants with predicted scores closest to 0, 1, and 2. We repeated this process to produce 10,000 sets of matches. From these sets, we retained only those where the score differences from 0, 1, and 2 were all below 0.1. Finally, we randomly chose four matches each from the White, Black, and Hispanic ethnicities, ensuring an equal distribution of two males and two females within each ethnic group.

Based on free-text, structured data, and imaging data, the hepatologist assigned each participant a classification of “No evidence of MASLD,” “Possible MASLD,” “Probable MASLD,” or “Definitive MASLD.” When possible, the hepatologist also identified their fibrosis or cirrhosis statuses. We tested the association between the independent variables (predicted MASLD score, age, ethnicity, gender) and the dependent variable (No evidence of MASLD < Possible MASLD < Probable MASLD < Definitive MASLD) using ordinal logistic regression via statsmodels (version 0.13.5).

### Supplemental Tables

**Supplemental Table 1:** ICD-10 codes used to define exclusionary diagnoses when selecting participants

| **Diagnosis** | **ICD-10 codes** |
| --- | --- |
| Exclusions: other liver diseases | |
| Alcoholic liver disease | K70 |
| Viral hepatitis | B16-B19 |
| Autoimmune liver disease (AIH, PBC, PSC) | K83.0A, K83.0F, K74.3, K75.4 |
| Hemochromatosis | E83.1 |
| Wilson disease | E83.0B |
| Alpha-1-antitrypsin deficiency | E88.0A, E88.0B |
| Budd-Chiari syndrome | I82.0, K76.5 |
| Chronic hepatitis, unspecified | K73.9, K73.2 |
| Secondary or unspecified biliary cirrhosis | K74.4, K74.5 |
| Exclusions: alcohol and drug use | |
| Codes associated with alcohol use disorder | F10 |
| Codes associated with somatic consequences of alcohol | E24.4, G62.1, I42.6, K29.2, G31.2, G72.1, K85.2, K86.0, T51.0, T51.9, Y57.3, X65, Z50.2, Z71.4, Z72.1 |
| Codes associated with drug use disorders except nicotine/caffeine | F11-F14, F16, F18, F19 |

**Abbreviations:** AIH: autoimmune hepatitis. PBC: primary biliary cholangitis. PSC: primary sclerosing cholangitis.

**Supplemental Table 2:** ICD-10 codes used to define MASLD outcomes

| **Diagnosis** | **ICD-10 codes** |
| --- | --- |
| Outcome: compensated cirrhosis | |
| Cirrhosis, compensated | K74.6 |
| Esophageal varices, not bleeding | I85.9, I98.2 |
| Gastric varices, not bleeding | I86.4 |
| Outcome: decompensated cirrhosis | |
| Esophageal varices, bleeding | I85.0, I98.3 |
| Ascites | R18 |
| Hepatorenal syndrome | K76.7 |
| Portal hypertension | K76.6 |
| Outcome: liver cancer | |
| Hepatocellular carcinoma | C22.0 |
| Liver cancer, unspecified | C22.9 |
| Outcome: liver transplantation | |
| Liver transplantation status | Z94.4 |

**Supplemental Table 3:** List of the 68 features used for machine learning models

| **Category** | **Features** |
| --- | --- |
| Demographics | 1. Age [years] 2. Gender 3. Ethnicity |
| Laboratory values | 1. Alanine transaminase (ALT) [U/L] 2. Alkaline phosphatase (ALK) [U/L] 3. Aspartate transaminase (AST) [U/L] 4. Albumin [mg/dL] 5. Basophil # [x 10^9^ cells/L] 6. Basophil % 7. Blood urea nitrogen (BUN) [mg/dL] 8. Calcium [mg/dL] 9. Cholesterol [mg/dL] 10. Creatinine [mg/dL] 11. Eosinophil # [x 10^9^ cells/L] 12. Eosinophil % 13. Glucose [mg/dL] 14. HDL cholesterol [mg/dL] 15. Hematocrit 16. Hemoglobin [g/dL] 17. Hemoglobin A1C (HbA1C) 18. LDL cholesterol [mg/dL] 19. Lymphocyte # [x 10^9^ cells/L] 20. Lymphocyte % 21. Mean corpuscular hemoglobin (MCH) [pg] 22. Mean corpuscular hemoglobin concentration (MCHC) [g/dL] 23. Mean corpuscular volume (MCV) [fL] 24. Mean platelet volume (MPV) [fL] 25. Monocyte # [x 10^9^ cells/L] 26. Monocyte % 27. Neutrophil # [x 10^9^ cells/L] 28. Neutrophil % 29. Platelet # [x 10^9^ cells/L] 30. Protein [g/dL] 31. Red blood cell # (RBC) [x 10^6^ cells/L] 32. Red cell distribution width (RDW) 33. Total bilirubin [mg/dL] 34. Triglycerides [mg/dL] 35. White blood cell # (WBC) [x 10^9^ cells/L] |
| Vital values | 1. Body mass index (BMI) 2. Height [cm] 3. Mean arterial pressure (MAP) [mm Hg] 4. Weight [kg] |
| Social values | 1. Alcohol intake frequency 2. Smoking status |
| Medications (ATC class) | 1. A02B (drugs for peptic ulcer and gastro-esophageal reflux disease) 2. A10B (blood glucose lowering drugs, excluding insulins) 3. C10A (lipid modifying agents, plain) 4. C10B (lipid modifying agents, combinations) |
| Elixhauser comorbidities | 1. Cardiac arrhythmia 2. Chronic pulmonary disease 3. Coagulopathy 4. Congestive heart failure 5. Deficiency anemia 6. Diabetes 7. Hypertension 8. Hypothyroidism 9. Leukemia/lymphoma 10. Liver disease 11. Metastatic cancer 12. Obesity 13. Other neurological disorders 14. Peptic ulcer disease 15. Peripheral vascular disorders 16. Pulmonary circulation disorders 17. Renal failure 18. Rheumatoid arthritis/collagen disorders 19. Solid tumor 20. Valvular disease |

For continuous features, units are listed as [unit]. Alcohol intake frequency is encoded as follows: {0: none, 1: less than monthly, 2: monthly, 3: weekly, 4: daily}. Smoking status, all medications, and all Elixhauser comorbidities are binary features, with 0 indicating no and 1 indicating yes.

**Supplemental Table 4:** List of ICD-10 codes used to define Elixhauser comorbidities

| **Elixhauser Comorbidity** | **ICD-10 Codes** |
| --- | --- |
| Congestive Heart Failure | I09.9, I11.0, I13.0, I13.2, I25.5, I42.0, I42.5, I42.6, I42.7, I42.8, I42.9, I43, I50, P29.0 |
| Cardiac Arrhythmia | I44.1, I44.2, I44.3, I45.6, I45.9, I47, I48, I49, R00.0, R00.1, R00.8, T82.1, Z45.0, Z95.0 |
| Valvular Disease | A52.0, I05, I06, I07, I08, I09.1, I09.8, I34, I35, I36, I37, I38, I39, Q23.0, Q23.1, Q23.2, Q23.3, Z95.2, Z95.3, Z95.4 |
| Pulmonary Circulation Disorders | I26, I27, I28.0, I28.8, I28.9 |
| Peripheral Vascular Disorders | I70, I71, I73.1, I73.8, I73.9, I77.1, I79.0, I79.2, K55.1, K55.8, K55.9, Z95.8, Z95.9 |
| Hypertension | I10, I11, I12, I13, I15 |
| Other Neurological Disorders | G10, G11, G12, G13, G20, G21, G22, G25.4, G25.5, G31.2, G31.8, G31.9, G32, G35, G36, G37, G40, G41, G93.1, G93.4, R47.0, R56 |
| Chronic Pulmonary Disease | I27.8, I27.9, J40, J41, J42, J43, J44, J45, J46, J47, J60, J61, J62, J63, J64, J65, J66, J67, J68.4, J70.1, J70.3 |
| Diabetes | E10, E11, E12, E13, E14 |
| Hypothyroidism | E00, E01, E02, E03, E89.0 |
| Renal Failure | I12.0, I13.1, N18, N19, N25.0, Z49.0, Z49.1, Z49.2, Z94.0, Z99.2 |
| Liver Disease | B18, I85, I86.4, I98.2, K70, K71.1, K71.3, K71.4, K71.5, K71.7, K72, K73, K74, K76.0, K76.2, K76.3, K76.4, K76.5, K76.6, K76.7, K76.8, K76.9, Z94.4 |
| Peptic Ulcer Disease | K25.7, K25.9, K26.7, K26.9, K27.7, K27.9, K28.7, K28.9 |
| Metastatic Cancer | C77, C78, C79, C80 |
| Solid Tumor | C0, C1, C2, C3, C4, C5, C6, C70, C71, C72, C73, C74, C75, C76, C97 |
| Rheumatoid Arthritis/Collagen Disorders | L94.0, L94.1, L94.3, M05, M06, M08, M12.0, M12.3, M30, M31.0, M31.1, M31.2, M31.3, M32, M33, M34, M35, M45, M46.1, M46.8, M46.9 |
| Obesity | E66 |
| Deficiency Anemia | D50.8, D50.9, D51, D52, D53 |
| Leukemia/Lymphoma | C8, C9 |
| Coagulopathy | D65, D66, D67, D68, D69.1, D69.3, D69.4, D69.5, D69.6 |

**Supplemental Table 5:** Parameters used to train LightGBM regression models

| **Parameter** | **Value** |
| --- | --- |
| objective | Regression |
| boosting_type | GOSS |
| num_iterations | 1500 |
| learning_rate | 0.005 |
| num_leaves | 63 |
| min_data_in_leaf | 20 |

**Supplemental Table 6:** Characteristics of UK Biobank participants

| **Feature** | **All participants** | **Training set** | **Prediction set** |
| --- | --- | --- | --- |
| MASLD score | | | |
| 0 | 406796 [96.9%] | 24157 [64.9%] | 382639 [100.0%] |
| 1 | 7595 [1.8%] | 7595 [20.4%] |  |
| 2 | 5128 [1.2%] | 5128 [13.8%] |  |
| 3 | 332 [0.1%] | 332 [0.9%] |  |
| Demographics | | | |
| Age | 58.6 [13.2] | 57.2 [12.3] | 58.8 [13.2] |
| Female | 230276 [54.8%] | 19538 [52.5%] | 210738 [55.1%] |
| Asian | 9564 [2.3%] | 590 [1.6%] | 8974 [2.3%] |
| Black | 6522 [1.6%] | 284 [0.8%] | 6238 [1.6%] |
| White | 403765 [96.2%] | 36338 [97.7%] | 367427 [96.0%] |
| Alcohol intake frequency | | | |
| Daily | 79301 [18.9%] | 5152 [13.8%] | 74149 [19.4%] |
| Weekly | 208759 [49.7%] | 20444 [54.9%] | 188315 [49.2%] |
| Monthly | 51155 [12.2%] | 6405 [17.2%] | 44750 [11.7%] |
| Less than monthly | 46971 [11.2%] | 2710 [7.3%] | 44261 [11.6%] |
| Smoking status | 184804 [44.0%] | 14204 [38.2%] | 170600 [44.6%] |
| Laboratory measurements | | | |
| ALK | 80.3 [28.5] | 78.5 [27.7] | 80.5 [28.6] |
| ALT | 20.1 [11.8] | 20.3 [12.5] | 20.0 [11.7] |
| AST | 24.3 [7.7] | 24.5 [7.9] | 24.3 [7.7] |
| Albumin | 4.5 [0.3] | 4.5 [0.3] | 4.5 [0.3] |
| BUN | 14.8 [4.6] | 14.7 [4.4] | 14.8 [4.6] |
| Basophil # | 0.0 [0.0] | 0.0 [0.0] | 0.0 [0.0] |
| Basophil % | 0.4 [0.4] | 0.4 [0.4] | 0.4 [0.4] |
| Calcium | 9.5 [0.4] | 9.5 [0.4] | 9.5 [0.4] |
| Cholesterol | 219.0 [58.7] | 219.6 [57.1] | 218.9 [58.8] |
| Creatinine | 0.8 [0.2] | 0.8 [0.2] | 0.8 [0.2] |
| Eosinophil # | 0.1 [0.1] | 0.1 [0.1] | 0.1 [0.1] |
| Eosinophil % | 2.1 [1.9] | 2.1 [1.9] | 2.1 [1.9] |
| Glucose | 88.9 [12.6] | 88.3 [12.1] | 88.9 [12.6] |
| HDL cholesterol | 54.4 [19.5] | 54.8 [19.7] | 54.4 [19.5] |
| HbA1C | 5.4 [0.5] | 5.4 [0.4] | 5.4 [0.5] |
| Hematocrit | 41.0 [4.8] | 41.1 [4.8] | 41.0 [4.8] |
| Hemoglobin | 14.1 [1.7] | 14.2 [1.7] | 14.1 [1.7] |
| LDL cholesterol | 136.3 [45.4] | 137.0 [44.4] | 136.3 [45.5] |
| Lymphocyte # | 1.9 [0.8] | 1.8 [0.7] | 1.9 [0.8] |
| Lymphocyte % | 28.6 [9.6] | 28.9 [9.4] | 28.5 [9.6] |
| MCH | 31.5 [2.0] | 31.5 [2.0] | 31.5 [2.0] |
| MCHC | 34.5 [1.2] | 34.5 [1.4] | 34.5 [1.2] |
| MCV | 91.2 [5.3] | 91.0 [5.2] | 91.2 [5.3] |
| MPV | 9.2 [1.4] | 9.2 [1.4] | 9.2 [1.3] |
| Monocyte # | 0.5 [0.2] | 0.4 [0.2] | 0.5 [0.2] |
| Monocyte % | 6.8 [2.6] | 6.8 [2.7] | 6.8 [2.6] |
| Neutrophil # | 4.0 [1.7] | 3.9 [1.6] | 4.0 [1.7] |
| Neutrophil % | 61.2 [10.8] | 60.9 [10.7] | 61.2 [10.9] |
| Platelet # | 247.9 [73.9] | 244.0 [72.5] | 248.0 [73.5] |
| Protein | 7.2 [0.5] | 7.2 [0.5] | 7.2 [0.5] |
| RBC | 4.5 [0.6] | 4.5 [0.5] | 4.5 [0.6] |
| RDW | 13.3 [0.9] | 13.3 [0.9] | 13.3 [1.0] |
| Total bilirubin | 0.5 [0.2] | 0.5 [0.2] | 0.5 [0.2] |
| Triglycerides | 131.1 [96.6] | 128.4 [95.6] | 131.3 [96.7] |
| WBC | 6.6 [2.2] | 6.5 [2.1] | 6.7 [2.2] |
| Vital measurements | | | |
| BMI | 26.7 [5.7] | 26.3 [5.5] | 26.7 [5.7] |
| Height | 168.0 [13.0] | 169.0 [13.7] | 168.0 [14.0] |
| MAP | 101.0 [16.0] | 99.7 [15.7] | 101.0 [16.3] |
| Weight | 76.3 [21.0] | 76.4 [20.8] | 76.3 [21.0] |
| Medications | | | |
| A02B | 51732 [12.3%] | 6491 [17.4%] | 45241 [11.8%] |
| A10B | 13732 [3.3%] | 1585 [4.3%] | 12147 [3.2%] |
| C10A | 19720 [4.7%] | 2508 [6.7%] | 17212 [4.5%] |
| C10B | 75013 [17.9%] | 8941 [24.0%] | 66072 [17.3%] |
| Elixhauser comorbidities | | | |
| Cardiac arrhythmia | 43637 [10.4%] | 3024 [8.1%] | 40613 [10.6%] |
| Chronic pulmonary  disease | 51672 [12.3%] | 3958 [10.6%] | 47714 [12.5%] |
| Coagulopathy | 4418 [1.1%] | 361 [1.0%] | 4057 [1.1%] |
| Congestive heart  failure | 13527 [3.2%] | 815 [2.2%] | 12712 [3.3%] |
| Deficiency anemia | 17737 [4.2%] | 1430 [3.8%] | 16307 [4.3%] |
| Diabetes | 33268 [7.9%] | 2855 [7.7%] | 30413 [7.9%] |
| Hypertension | 120740 [28.8%] | 9054 [24.3%] | 111686 [29.2%] |
| Hypothyroidism | 24758 [5.9%] | 1940 [5.2%] | 22818 [6.0%] |
| Leukemia and  lymphoma | 9145 [2.2%] | 584 [1.6%] | 8561 [2.2%] |
| Liver disease | 3809 [0.9%] | 505 [1.4%] | 3304 [0.9%] |
| Metastatic cancer | 18214 [4.3%] | 877 [2.4%] | 17337 [4.5%] |
| Obesity | 27432 [6.5%] | 2507 [6.7%] | 24925 [6.5%] |
| Other neurological  disorders | 15092 [3.6%] | 859 [2.3%] | 14233 [3.7%] |
| Peptic ulcer disease | 9200 [2.2%] | 808 [2.2%] | 8392 [2.2%] |
| Peripheral vascular  disorders | 12337 [2.9%] | 724 [1.9%] | 11613 [3.0%] |
| Pulmonary circulation  disorders | 8596 [2.0%] | 578 [1.6%] | 8018 [2.1%] |
| Renal failure | 17319 [4.1%] | 1217 [3.3%] | 16102 [4.2%] |
| Rheumatoid arthritis  and collagen disorders | 15059 [3.6%] | 1132 [3.0%] | 13927 [3.6%] |
| Solid tumor | 70443 [16.8%] | 5138 [13.8%] | 65305 [17.1%] |
| Valvular disease | 16383 [3.9%] | 1069 [2.9%] | 15314 [4.0%] |

Values are expressed as either median with [interquartile range] or count with [percent of total participants]. **Abbreviations:** ALK: alkaline phosphatase. ALT: alanine aminotransferase. AST: aspartate aminotransferase. BUN: blood urea nitrogen. HbA1C: hemoglobin A1C. MCH: mean corpuscular hemoglobin. MCHC: mean corpuscular hemoglobin concentration. MCV: mean corpuscular volume. MPV: mean platelet volume. RBC: red blood cell count. RDW: red blood cell distribution width. BMI: body mass index. MAP: mean arterial pressure.

**Supplemental Table 7:** Characteristics of participants in external datasets

| **Feature** | **MSDW** | **Bio*Me*** | ***All of Us*** |
| --- | --- | --- | --- |
| MASLD score | | | |
| 0 | 109504 [99.0%] | 18114 [96.5%] | 81382 [93.2%] |
| 1 | 543 [0.5%] | 381 [2.0%] | 3494 [4.0%] |
| 2 | 322 [0.3%] | 231 [1.2%] | 2058 [2.4%] |
| 3 | 238 [0.2%] | 45 [0.2%] | 374 [0.4%] |
| Demographics | | | |
| Age | 55.6 [34.4] | 59.3 [27.3] | 59.1 [27.8] |
| Female | 64730 [57.5%] | 17780 [62.0%] | 72690 [65.2%] |
| Asian | 11202 [10.0%] | 1056 [3.7%] | 3269 [2.9%] |
| Black | 23414 [20.8%] | 6455 [22.5%] | 15987 [14.3%] |
| White | 77184 [68.6%] | 11263 [39.3%] | 68200 [61.1%] |
| Other races* | 714 [0.6%] | 9899 [34.5%] | 24082 [21.6%] |
| Alcohol intake frequency | | | |
| Daily | 3822 [3.4%] | 1109 [3.9%] | 77151 [69.2%] |
| Weekly | 391 [0.3%] | 4829 [16.8%] | 266 [0.2%] |
| Monthly | 582 [0.5%] | 3713 [12.9%] | 406 [0.4%] |
| Less than monthly | 46278 [41.1%] | 6552 [22.9%] | 20774 [18.6%] |
| Smoking status | 35226 [31.3%] | 10784 [37.6%] | 62010 [55.6%] |
| Laboratory measurements | | | |
| ALK | 70.0 [30.0] | 75.0 [31.5] | 73.5 [30.0] |
| ALT | 18.5 [12.0] | 19.0 [12.0] | 20.0 [12.0] |
| AST | 21.0 [8.5] | 21.0 [8.0] | 21.0 [9.0] |
| Albumin | 4.2 [0.8] | 4.0 [0.6] | 4.0 [0.8] |
| BUN | 14.5 [7.0] | 15.3 [5.9] | 14.5 [6.5] |
| Basophil # | 0.0 [0.1] | 0.0 [0.0] | 0.0 [0.0] |
| Basophil % | 0.5 [0.7] | 0.5 [0.4] | 0.6 [0.5] |
| Calcium | 9.4 [0.7] | 9.3 [0.4] | 9.3 [0.6] |
| Cholesterol | 178.7 [38.5] | 179.3 [44.1] | 177.0 [38.0] |
| Creatinine | 0.8 [0.3] | 1.6 [1.4] | 0.8 [0.3] |
| Eosinophil # | 0.1 [0.1] | 0.1 [0.1] | 0.1 [0.1] |
| Eosinophil % | 1.9 [1.9] | 1.9 [1.9] | 2.0 [1.9] |
| Glucose | 95.0 [21.0] | 93.5 [23.5] | 99.5 [23.0] |
| HDL cholesterol | 54.5 [19.8] | 53.0 [19.8] | 52.7 [18.0] |
| HbA1C | 5.6 [0.7] | 5.7 [0.9] | 5.6 [0.8] |
| Hematocrit | 40.0 [6.4] | 39.4 [6.1] | 39.8 [5.6] |
| Hemoglobin | 13.3 [2.4] | 13.0 [2.2] | 13.2 [2.0] |
| LDL cholesterol | 99.8 [30.2] | 99.3 [38.8] | 101.6 [37.8] |
| Lymphocyte # | 1.7 [0.9] | 1.8 [0.8] | 1.8 [0.9] |
| Lymphocyte % | 26.4 [14.0] | 27.0 [13.1] | 26.3 [12.4] |
| MCH | 30.1 [2.5] | 29.8 [2.7] | 29.9 [2.6] |
| MCHC | 33.3 [1.2] | 33.1 [1.0] | 33.3 [1.4] |
| MCV | 90.2 [6.5] | 89.6 [6.9] | 89.8 [6.5] |
| MPV | 8.7 [1.5] | 8.8 [1.3] | 9.8 [1.6] |
| Monocyte # | 0.5 [0.3] | 0.5 [0.2] | 0.6 [0.3] |
| Monocyte % | 7.6 [2.7] | 7.4 [2.6] | 7.9 [2.6] |
| Neutrophil # | 4.3 [2.8] | 4.2 [2.4] | 4.3 [2.7] |
| Neutrophil % | 62.5 [14.6] | 62.1 [14.0] | 62.0 [13.0] |
| Platelet # | 239.0 [86.0] | 237.0 [82.5] | 242.5 [83.5] |
| Protein | 7.0 [0.7] | 7.1 [0.4] | 7.1 [0.6] |
| RBC | 4.4 [0.8] | 4.4 [0.7] | 4.5 [0.7] |
| RDW | 13.2 [1.6] | 13.9 [1.6] | 13.5 [1.4] |
| Total bilirubin | 0.5 [0.4] | 0.4 [0.3] | 0.5 [0.3] |
| Triglycerides | 102.4 [61.7] | 111.5 [67.0] | 114.8 [60.5] |
| WBC | 7.0 [3.0] | 6.9 [2.8] | 7.0 [3.0] |
| Vital measurements | | | |
| BMI | 26.1 [7.5] | 27.7 [8.2] | 29.0 [9.4] |
| Height | 167.6 [15.2] | 165.1 [15.2] | 167.1 [15.2] |
| MAP | 96.5 [12.5] | 91.2 [11.5] | 92.7 [12.7] |
| Weight | 74.4 [25.9] | 77.1 [25.7] | 81.6 [29.0] |
| Medications | | | |
| A02B | 37977 [33.8%] | 14518 [50.6%] | 62639 [56.2%] |
| A10B | 9096 [8.1%] | 5158 [18.0%] | 20880 [18.7%] |
| C10A | 27900 [24.8%] | 12413 [43.3%] | 47276 [42.4%] |
| C10B | 213 [0.2%] | 144 [0.5%] | 420 [0.4%] |
| Elixhauser comorbidities | | | |
| Cardiac arrhythmia | 16327 [14.5%] | 5320 [18.6%] | 33245 [29.8%] |
| Chronic pulmonary  disease | 18406 [16.4%] | 7737 [27.0%] | 36231 [32.5%] |
| Coagulopathy | 1532 [1.4%] | 1617 [5.6%] | 8872 [8.0%] |
| Congestive heart  failure | 9735 [8.7%] | 3015 [10.5%] | 12191 [10.9%] |
| Deficiency anemia | 6257 [5.6%] | 3425 [11.9%] | 15997 [14.3%] |
| Diabetes | 21720 [19.3%] | 7806 [27.2%] | 30212 [27.1%] |
| Hypertension | 13953 [12.4%] | 14621 [51.0%] | 56883 [51.0%] |
| Hypothyroidism | 4641 [4.1%] | 4227 [14.7%] | 21296 [19.1%] |
| Leukemia and  lymphoma | 6601 [5.9%] | 879 [3.1%] | 6091 [5.5%] |
| Liver disease | 1726 [1.5%] | 683 [2.4%] | 9002 [8.1%] |
| Metastatic cancer | 2247 [2.0%] | 552 [1.9%] | 4215 [3.8%] |
| Obesity | 23324 [20.7%] | 10947 [38.2%] | 39792 [35.7%] |
| Other neurological  disorders | 5718 [5.1%] | 2275 [7.9%] | 12277 [11.0%] |
| Peptic ulcer disease | 1000 [0.9%] | 700 [2.4%] | 3852 [3.5%] |
| Peripheral vascular  disorders | 9183 [8.2%] | 4190 [14.6%] | 13657 [12.2%] |
| Pulmonary circulation  disorders | 4247 [3.8%] | 1340 [4.7%] | 7135 [6.4%] |
| Renal failure | 7207 [6.4%] | 4174 [14.6%] | 13555 [12.2%] |
| Rheumatoid arthritis  and collagen disorders | 4631 [4.1%] | 2513 [8.8%] | 19321 [17.3%] |
| Solid tumor | 34228 [30.4%] | 3997 [13.9%] | 24115 [21.6%] |
| Valvular disease | 10051 [8.9%] | 2938 [10.2%] | 15478 [13.9%] |

Values are expressed as either median with [interquartile range] or count with [percent of total participants]. *: In MSDW and All of Us, most Hispanic participants also selected a self-identification as either White or Black, in which case they were included under those categories; in BioMe, all Hispanic participants were included under “Other races” as a secondary self-identification was not available. **Abbreviations:** ALK: alkaline phosphatase. ALT: alanine aminotransferase. AST: aspartate aminotransferase. BUN: blood urea nitrogen. HbA1C: hemoglobin A1C. MCH: mean corpuscular hemoglobin. MCHC: mean corpuscular hemoglobin concentration. MCV: mean corpuscular volume. MPV: mean platelet volume. RBC: red blood cell count. RDW: red blood cell distribution width. BMI: body mass index. MAP: mean arterial pressure.

**Supplemental Table 8:** Regression metrics for the UK Biobank holdout set

| **Dataset** | **RMSE** | **MAE** | **R^2^** | **Spearman’s ρ** |
| --- | --- | --- | --- | --- |
| MASLD and MetALD | 0.61 (0.61-0.61) | 0.45 (0.45-0.45) | 0.36 (0.36-0.36) | 0.56 (0.56-0.56) |
| MetALD only | 0.59 (0.59-0.59) | 0.43 (0.43-0.43) | 0.36 (0.36-0.36) | 0.57 (0.56-0.57) |

**Abbreviations:** RMSE: root-mean-square error. MAE: mean absolute error.

**Supplemental Table 9:** Classification performance of the hepatic steatosis index (HSI)

| **Task** | **Dataset** | **AUROC** | **AUPRC** | **Proportion** |
| --- | --- | --- | --- | --- |
| Holdout (MASLD and MetALD) | | | | |
| MASLD diagnosis | UK Biobank | 0.80 (0.80-0.80) | 0.68 (0.68-0.68) | 0.35 |
| Fibrosis identification |  | 0.61 (0.60-0.62) | 0.02 (0.02-0.02) | 0.01 |
| Holdout (MASLD only) | | | | |
| MASLD diagnosis | UK Biobank | 0.81 (0.81-0.81) | 0.68 (0.68-0.68) | 0.34 |
| Fibrosis identification |  | 0.65 (0.63-0.67) | 0.02 (0.02-0.02) | 0.01 |

**Abbreviations:** AUROC: area under the precision recall curve. AUPRC: area under the precision recall curve.

**Supplemental Table 10:** Classification performance of the FIB-4 index

| **Task** | **Dataset** | **AUROC** | **AUPRC** | **Proportion** |
| --- | --- | --- | --- | --- |
| Holdout (MASLD and MetALD) | | | | |
| MASLD diagnosis | UK Biobank | 0.45 (0.45-0.45) | 0.33 (0.33-0.33) | 0.35 |
| Fibrosis identification |  | 0.99 (0.99-0.99) | 0.42 (0.41-0.43) | 0.01 |
| Holdout (MASLD only) | | | | |
| MASLD diagnosis | UK Biobank | 0.44 (0.44-0.44) | 0.31 (0.31-0.31) | 0.34 |
| Fibrosis identification |  | 0.99 (0.99-0.99) | 0.44 (0.42-0.46) | 0.01 |

**Abbreviations:** AUROC: area under the precision recall curve. AUPRC: area under the precision recall curve.

**Supplemental Table 11:** Additional classification metrics for MASLD diagnosis in the holdout set

| **Threshold** | **Sensitivity** | **Specificity** | **PPV** | **NPV** |
| --- | --- | --- | --- | --- |
| Holdout (MASLD and MetALD) | | | | |
| MASLD score | | | | |
| 0.25 | 0.92 (0.92-0.92) | 0.50 (0.50-0.50) | 0.50 (0.50-0.50) | 0.92 (0.92-0.92) |
| 0.50 | 0.77 (0.77-0.77) | 0.74 (0.74-0.74) | 0.61 (0.61-0.61) | 0.85 (0.85-0.85) |
| 0.75 | 0.53 (0.53-0.53) | 0.88 (0.88-0.88) | 0.71 (0.71-0.71) | 0.77 (0.77-0.77) |
| 1.00 | 0.30 (0.30-0.30) | 0.95 (0.95-0.95) | 0.78 (0.78-0.78) | 0.71 (0.71-0.71) |
| 1.25 | 0.16 (0.16-0.16) | 0.98 (0.98-0.98) | 0.84 (0.84-0.84) | 0.68 (0.68-0.68) |
| 1.50 | 0.08 (0.08-0.08) | 0.99 (0.99-0.99) | 0.89 (0.88-0.90) | 0.67 (0.67-0.67) |
| 1.75 | 0.04 (0.04-0.04) | 1.00 (1.00-1.00) | 0.93 (0.92-0.94) | 0.66 (0.66-0.66) |
| 2.00 | 0.02 (0.02-0.02) | 1.00 (1.00-1.00) | 0.94 (0.93-0.95) | 0.65 (0.65-0.65) |
| FIB-4 score |  |  |  |  |
| 1.30 | 0.42 (0.42-0.42) | 0.52 (0.52-0.52) | 0.32 (0.32-0.32) | 0.62 (0.62-0.62) |
| 2.67 | 0.03 (0.03-0.03) | 0.98 (0.98-0.98) | 0.37 (0.36-0.38) | 0.65 (0.65-0.65) |
| Holdout (MASLD only) | | | | |
| MASLD score | | | | |
| 0.25 | 0.92 (0.92-0.92) | 0.52 (0.52-0.52) | 0.50 (0.50-0.50) | 0.93 (0.93-0.93) |
| 0.50 | 0.76 (0.76-0.76) | 0.75 (0.75-0.75) | 0.61 (0.61-0.61) | 0.86 (0.86-0.86) |
| 0.75 | 0.53 (0.53-0.53) | 0.89 (0.89-0.89) | 0.72 (0.72-0.72) | 0.79 (0.79-0.79) |
| 1.00 | 0.29 (0.29-0.29) | 0.96 (0.96-0.96) | 0.79 (0.79-0.79) | 0.72 (0.72-0.72) |
| 1.25 | 0.14 (0.14-0.14) | 0.99 (0.99-0.99) | 0.84 (0.84-0.84) | 0.69 (0.69-0.69) |
| 1.50 | 0.07 (0.07-0.07) | 1.00 (1.00-1.00) | 0.89 (0.88-0.90) | 0.68 (0.68-0.68) |
| 1.75 | 0.03 (0.03-0.03) | 1.00 (1.00-1.00) | 0.94 (0.93-0.95) | 0.67 (0.67-0.67) |
| 2.00 | 0.01 (0.01-0.01) | 1.00 (1.00-1.00) | 0.96 (0.95-0.97) | 0.66 (0.66-0.66) |
| FIB-4 score |  |  |  |  |
| 1.30 | 0.40 (0.40-0.40) | 0.51 (0.51-0.51) | 0.30 (0.30-0.30) | 0.62 (0.62-0.62) |
| 2.67 | 0.02 (0.02-0.02) | 0.98 (0.98-0.98) | 0.34 (0.33-0.35) | 0.66 (0.66-0.66) |

**Abbreviations:** PPV: positive predictive value. NPV: negative predictive value.

**Supplemental Table 12:** Additional classification metrics for fibrosis identification in the holdout set

| **Threshold** | **Sensitivity** | **Specificity** | **PPV** | **NPV** |
| --- | --- | --- | --- | --- |
| Holdout (MASLD and MetALD) | | | | |
| MASLD score | | | | |
| 0.25 | 0.99 (0.99-0.99) | 0.35 (0.35-0.35) | 0.01 (0.01-0.01) | 1.00 (1.00-1.00) |
| 0.50 | 0.93 (0.92-0.94) | 0.57 (0.57-0.57) | 0.02 (0.02-0.02) | 1.00 (1.00-1.00) |
| 0.75 | 0.86 (0.85-0.87) | 0.75 (0.75-0.75) | 0.03 (0.03-0.03) | 1.00 (1.00-1.00) |
| 1.00 | 0.78 (0.76-0.80) | 0.87 (0.87-0.87) | 0.05 (0.05-0.05) | 1.00 (1.00-1.00) |
| 1.25 | 0.67 (0.64-0.70) | 0.94 (0.94-0.94) | 0.09 (0.09-0.09) | 1.00 (1.00-1.00) |
| 1.50 | 0.56 (0.54-0.58) | 0.97 (0.97-0.97) | 0.16 (0.15-0.17) | 1.00 (1.00-1.00) |
| 1.75 | 0.46 (0.44-0.48) | 0.99 (0.99-0.99) | 0.29 (0.28-0.30) | 1.00 (1.00-1.00) |
| 2.00 | 0.32 (0.30-0.34) | 1.00 (1.00-1.00) | 0.48 (0.45-0.51) | 0.99 (0.99-0.99) |
| FIB-4 score |  |  |  |  |
| 1.30 | 1.00 (1.00-1.00) | 0.54 (0.54-0.54) | 0.02 (0.02-0.02) | 1.00 (1.00-1.00) |
| 2.67 | 1.00 (1.00-1.00) | 0.98 (0.98-0.98) | 0.37 (0.36-0.38) | 1.00 (1.00-1.00) |
| Holdout (MASLD only) | | | | |
| MASLD score | | | | |
| 0.25 | 0.98 (0.97-0.99) | 0.37 (0.37-0.37) | 0.01 (0.01-0.01) | 1.00 (1.00-1.00) |
| 0.50 | 0.93 (0.92-0.94) | 0.58 (0.58-0.58) | 0.02 (0.02-0.02) | 1.00 (1.00-1.00) |
| 0.75 | 0.84 (0.82-0.86) | 0.76 (0.76-0.76) | 0.03 (0.03-0.03) | 1.00 (1.00-1.00) |
| 1.00 | 0.77 (0.74-0.80) | 0.88 (0.88-0.88) | 0.05 (0.05-0.05) | 1.00 (1.00-1.00) |
| 1.25 | 0.68 (0.65-0.71) | 0.95 (0.95-0.95) | 0.09 (0.09-0.09) | 1.00 (1.00-1.00) |
| 1.50 | 0.56 (0.53-0.59) | 0.98 (0.98-0.98) | 0.16 (0.15-0.17) | 1.00 (1.00-1.00) |
| 1.75 | 0.48 (0.45-0.51) | 0.99 (0.99-0.99) | 0.30 (0.29-0.31) | 1.00 (1.00-1.00) |
| 2.00 | 0.35 (0.32-0.38) | 1.00 (1.00-1.00) | 0.51 (0.49-0.53) | 0.99 (0.99-0.99) |
| FIB-4 score |  |  |  |  |
| 1.30 | 1.00 (1.00-1.00) | 0.54 (0.54-0.54) | 0.02 (0.02-0.02) | 1.00 (1.00-1.00) |
| 2.67 | 1.00 (1.00-1.00) | 0.98 (0.98-0.98) | 0.34 (0.33-0.35) | 1.00 (1.00-1.00) |

**Abbreviations:** PPV: positive predictive value. NPV: negative predictive value.

**Supplemental Table 13:** Classification metrics for models evaluated on subsets of the UK Biobank holdout set.

| **Task** | **Subset** | **AUROC** | **AUPRC** | **Proportion** |
| --- | --- | --- | --- | --- |
| MASLD and MetALD | | | | |
| MASLD diagnosis | All | 0.83 (0.83-0.83) | 0.71 (0.71-0.71) | 0.35 |
|  | Asian | 0.81 (0.8-0.82) | 0.73 (0.72-0.74) |  |
|  | Black | 0.83 (0.82-0.84) | 0.71 (0.70-0.72) |  |
|  | White | 0.83 (0.83-0.83) | 0.71 (0.71-0.71) |  |
|  | Age 40-69 | 0.83 (0.83-0.83) | 0.72 (0.72-0.72) |  |
|  | Age > 69 | 0.77 (0.76-0.78) | 0.58 (0.57-0.59) |  |
|  | Female | 0.86 (0.86-0.86) | 0.77 (0.77-0.77) |  |
|  | Male | 0.78 (0.78-0.78) | 0.64 (0.64-0.64) |  |
|  | 18.5 ≥ BMI < 25 | 0.79 (0.79-0.79) | 0.67 (0.66-0.68) |  |
|  | 25 ≥ BMI < 30 | 0.79 (0.79-0.79) | 0.67 (0.66-0.68) |  |
|  | BMI > 30 | 0.71 (0.71-0.71) | 0.56 (0.56-0.56) |  |
| Fibrosis identification | All | 0.91 (0.90-0.92) | 0.36 (0.34-0.38) | 0.01 |
|  | Asian | Insufficient sample size | |  |
|  | Black | 0.97 (0.97-0.97) | 0.20 (0.18-0.22) |  |
|  | White | 0.91 (0.91-0.91) | 0.36 (0.36-0.36) |  |
|  | Age 40-69 | 0.91 (0.91-0.91) | 0.37 (0.37-0.37) |  |
|  | Age > 69 | 0.87 (0.81-0.93) | 0.34 (0.21-0.47) |  |
|  | Female | 0.91 (0.91-0.91) | 0.46 (0.46-0.46) |  |
|  | Male | 0.89 (0.88-0.90) | 0.30 (0.28-0.32) |  |
|  | 18.5 ≥ BMI < 25 | 0.85 (0.85-0.85) | 0.08 (0.07-0.09) |  |
|  | 25 ≥ BMI < 30 | 0.85 (0.85-0.85) | 0.08 (0.07-0.09) |  |
|  | BMI > 30 | 0.95 (0.94-0.96) | 0.43 (0.37-0.49) |  |
| MASLD only | | | | |
| MASLD diagnosis | All | 0.84 (0.84-0.84) | 0.71 (0.71-0.71) | 0.34 |
|  | Asian | 0.80 (0.79-0.81) | 0.69 (0.68-0.70) |  |
|  | Black | 0.84 (0.84-0.84) | 0.73 (0.72-0.74) |  |
|  | White | 0.84 (0.84-0.84) | 0.71 (0.71-0.71) |  |
|  | Other | 0.84 (0.84-0.84) | 0.72 (0.72-0.72) |  |
|  | Age < 40 | 0.81 (0.80-0.82) | 0.62 (0.60-0.64) |  |
|  | Age 40-69 | 0.87 (0.87-0.87) | 0.77 (0.77-0.77) |  |
|  | Age > 69 | 0.80 (0.80-0.80) | 0.65 (0.65-0.65) |  |
|  | Female | 0.80 (0.80-0.80) | 0.68 (0.67-0.69) |  |
|  | Male | 0.80 (0.80-0.80) | 0.68 (0.67-0.69) |  |
|  | 18.5 ≥ BMI < 25 | 0.71 (0.7-0.72) | 0.55 (0.54-0.56) |  |
|  | 25 ≥ BMI < 30 | 0.80 (0.79-0.81) | 0.69 (0.68-0.70) |  |
|  | BMI > 30 | 0.84 (0.84-0.84) | 0.73 (0.72-0.74) |  |
| Fibrosis identification | All | 0.90 (0.89-0.91) | 0.40 (0.38-0.42) | 0.01 |
|  | Asian | Insufficient sample size | |  |
|  | Black | 0.97 (0.97-0.97) | 0.26 (0.24-0.28) |  |
|  | White | 0.90 (0.90-0.90) | 0.39 (0.39-0.39) |  |
|  | Other | 0.91 (0.91-0.91) | 0.39 (0.39-0.39) |  |
|  | Age < 40 | 0.81 (0.73-0.89) | 0.28 (0.15-0.41) |  |
|  | Age 40-69 | 0.91 (0.91-0.91) | 0.48 (0.48-0.48) |  |
|  | Age > 69 | 0.88 (0.87-0.89) | 0.33 (0.31-0.35) |  |
|  | Female | 0.84 (0.84-0.84) | 0.05 (0.05-0.05) |  |
|  | Male | 0.84 (0.84-0.84) | 0.05 (0.05-0.05) |  |
|  | 18.5 ≥ BMI < 25 | 0.96 (0.94-0.98) | 0.53 (0.49-0.57) |  |
|  | 25 ≥ BMI < 30 | 0.97 (0.97-0.97) | 0.26 (0.24-0.28) |  |
|  | BMI > 30 | 0.90 (0.90-0.90) | 0.39 (0.39-0.39) |  |

**Abbreviations:** AUROC: area under the precision recall curve. AUPRC: area under the precision recall curve.

**Supplemental Table 14:** Classification metrics for models evaluated on subsets of the MSDW dataset

| **Task** | **Subset** | **AUROC** | **AUPRC** | **Proportion** |
| --- | --- | --- | --- | --- |
| MASLD and MetALD | | | | |
| MASLD diagnosis | All | 0.79 (0.78-0.80) | 0.62 (0.61-0.63) | 0.35 |
|  | Asian | 0.84 (0.83-0.85) | 0.70 (0.68-0.72) |  |
|  | Black | 0.77 (0.76-0.78) | 0.59 (0.57-0.61) |  |
|  | White | 0.80 (0.80-0.80) | 0.63 (0.62-0.64) |  |
|  | Other | 0.81 (0.77-0.85) | 0.70 (0.64-0.76) |  |
|  | Age < 40 | 0.92 (0.90-0.94) | 0.86 (0.83-0.89) |  |
|  | Age 40-69 | 0.80 (0.80-0.80) | 0.64 (0.63-0.65) |  |
|  | Age > 69 | 0.66 (0.64-0.68) | 0.45 (0.43-0.47) |  |
|  | Female | 0.82 (0.81-0.83) | 0.66 (0.65-0.67) |  |
|  | Male | 0.77 (0.76-0.78) | 0.58 (0.57-0.59) |  |
|  | 18.5 ≥ BMI < 25 | 0.81 (0.80-0.82) | 0.62 (0.59-0.65) |  |
|  | 25 ≥ BMI < 30 | 0.81 (0.80-0.82) | 0.62 (0.59-0.65) |  |
|  | BMI > 30 | 0.68 (0.67-0.69) | 0.50 (0.49-0.51) |  |
| Fibrosis identification | All | 0.89 (0.87-0.91) | 0.37 (0.29-0.45) | 0.01 |
|  | Asian | Insufficient sample size | |  |
|  | Black | 0.87 (0.78-0.96) | 0.53 (0.33-0.73) |  |
|  | White | 0.90 (0.86-0.94) | 0.34 (0.25-0.43) |  |
|  | Other | Insufficient sample size | |  |
|  | Age < 40 | 0.67 (0.51-0.83) | 0.16 (0.01-0.31) |  |
|  | Age 40-69 | 0.87 (0.83-0.91) | 0.31 (0.20-0.42) |  |
|  | Age > 69 | 0.79 (0.68-0.9) | 0.29 (0.13-0.45) |  |
|  | Female | 0.90 (0.86-0.94) | 0.35 (0.22-0.48) |  |
|  | Male | 0.89 (0.86-0.92) | 0.26 (0.19-0.33) |  |
|  | 18.5 ≥ BMI < 25 | 0.81 (0.71-0.91) | 0.25 (0.07-0.43) |  |
|  | 25 ≥ BMI < 30 | 0.81 (0.71-0.91) | 0.25 (0.07-0.43) |  |
|  | BMI > 30 | 0.96 (0.94-0.98) | 0.35 (0.26-0.44) |  |
| MASLD only | | | | |
| MASLD diagnosis | All | 0.80 (0.80-0.80) | 0.61 (0.60-0.62) | 0.34 |
|  | Asian | 0.82 (0.80-0.84) | 0.66 (0.63-0.69) |  |
|  | Black | 0.76 (0.75-0.77) | 0.57 (0.55-0.59) |  |
|  | White | 0.80 (0.79-0.81) | 0.62 (0.61-0.63) |  |
|  | Other | 0.80 (0.74-0.86) | 0.70 (0.60-0.80) |  |
|  | Age < 40 | 0.90 (0.87-0.93) | 0.84 (0.79-0.89) |  |
|  | Age 40-69 | 0.79 (0.79-0.79) | 0.62 (0.61-0.63) |  |
|  | Age > 69 | 0.65 (0.62-0.68) | 0.44 (0.42-0.46) |  |
|  | Female | 0.81 (0.80-0.82) | 0.64 (0.63-0.65) |  |
|  | Male | 0.76 (0.75-0.77) | 0.56 (0.55-0.57) |  |
|  | 18.5 ≥ BMI < 25 | 0.79 (0.76-0.82) | 0.58 (0.53-0.63) |  |
|  | 25 ≥ BMI < 30 | 0.79 (0.76-0.82) | 0.58 (0.53-0.63) |  |
|  | BMI > 30 | 0.67 (0.67-0.67) | 0.47 (0.46-0.48) |  |
| Fibrosis identification | All | 0.91 (0.89-0.93) | 0.38 (0.31-0.45) | 0.01 |
|  | Asian | Insufficient sample size | |  |
|  | Black | 0.91 (0.81-1.01) | 0.54 (0.34-0.74) |  |
|  | White | 0.91 (0.89-0.93) | 0.32 (0.23-0.41) |  |
|  | Other | Insufficient sample size | |  |
|  | Age < 40 | 0.60 (0.40-0.80) | 0.32 (0.00-0.66) |  |
|  | Age 40-69 | 0.92 (0.90-0.94) | 0.37 (0.24-0.50) |  |
|  | Age > 69 | 0.87 (0.78-0.96) | 0.36 (0.19-0.53) |  |
|  | Female | 0.89 (0.85-0.93) | 0.37 (0.27-0.47) |  |
|  | Male | 0.90 (0.85-0.95) | 0.31 (0.19-0.43) |  |
|  | 18.5 ≥ BMI < 25 | 0.85 (0.73-0.97) | 0.35 (0.02-0.68) |  |
|  | 25 ≥ BMI < 30 | 0.85 (0.73-0.97) | 0.35 (0.02-0.68) |  |
|  | BMI > 30 | 0.96 (0.94-0.98) | 0.34 (0.24-0.44) |  |

**Abbreviations:** AUROC: area under the precision recall curve. AUPRC: area under the precision recall curve.

**Supplemental Table 15:** Classification metrics for models evaluated on subsets of the *All of Us* dataset

| **Task** | **Subset** | **AUROC** | **AUPRC** | **Proportion** |
| --- | --- | --- | --- | --- |
| MASLD and MetALD | | | | |
| MASLD diagnosis | All | 0.74 (0.74-0.74) | 0.55 (0.55-0.55) | 0.35 |
|  | Asian | 0.82 (0.81-0.83) | 0.66 (0.64-0.68) |  |
|  | Black | 0.73 (0.72-0.74) | 0.54 (0.53-0.55) |  |
|  | White | 0.74 (0.74-0.74) | 0.55 (0.55-0.55) |  |
|  | Other | 0.75 (0.75-0.75) | 0.57 (0.56-0.58) |  |
|  | Age < 40 | 0.85 (0.84-0.86) | 0.76 (0.74-0.78) |  |
|  | Age 40-69 | 0.75 (0.75-0.75) | 0.58 (0.58-0.58) |  |
|  | Age > 69 | 0.66 (0.65-0.67) | 0.44 (0.43-0.45) |  |
|  | Female | 0.77 (0.77-0.77) | 0.60 (0.60-0.60) |  |
|  | Male | 0.69 (0.69-0.69) | 0.49 (0.49-0.49) |  |
|  | 18.5 ≥ BMI < 25 | 0.76 (0.76-0.76) | 0.60 (0.59-0.61) |  |
|  | 25 ≥ BMI < 30 | 0.76 (0.76-0.76) | 0.60 (0.59-0.61) |  |
|  | BMI > 30 | 0.66 (0.66-0.66) | 0.48 (0.48-0.48) |  |
| Fibrosis identification | All | 0.89 (0.89-0.89) | 0.24 (0.22-0.26) | 0.01 |
|  | Asian | 0.77 (0.69-0.85) | 0.27 (0.18-0.36) |  |
|  | Black | 0.84 (0.82-0.86) | 0.18 (0.15-0.21) |  |
|  | White | 0.90 (0.90-0.90) | 0.26 (0.24-0.28) |  |
|  | Other | 0.92 (0.90-0.94) | 0.28 (0.24-0.32) |  |
|  | Age < 40 | 0.89 (0.83-0.95) | 0.11 (0.08-0.14) |  |
|  | Age 40-69 | 0.92 (0.91-0.93) | 0.34 (0.32-0.36) |  |
|  | Age > 69 | 0.84 (0.80-0.88) | 0.17 (0.15-0.19) |  |
|  | Female | 0.89 (0.88-0.90) | 0.28 (0.26-0.30) |  |
|  | Male | 0.88 (0.87-0.89) | 0.24 (0.21-0.27) |  |
|  | 18.5 ≥ BMI < 25 | 0.87 (0.85-0.89) | 0.08 (0.06-0.10) |  |
|  | 25 ≥ BMI < 30 | 0.87 (0.85-0.89) | 0.08 (0.06-0.10) |  |
|  | BMI > 30 | 0.94 (0.94-0.94) | 0.28 (0.26-0.30) |  |
| MASLD only | | | | |
| MASLD diagnosis | All | 0.74 (0.74-0.74) | 0.54 (0.54-0.54) | 0.34 |
|  | Asian | 0.83 (0.82-0.84) | 0.67 (0.65-0.69) |  |
|  | Black | 0.73 (0.72-0.74) | 0.53 (0.52-0.54) |  |
|  | White | 0.75 (0.75-0.75) | 0.54 (0.54-0.54) |  |
|  | Other | 0.74 (0.74-0.74) | 0.54 (0.53-0.55) |  |
|  | Age < 40 | 0.86 (0.85-0.87) | 0.77 (0.74-0.80) |  |
|  | Age 40-69 | 0.76 (0.76-0.76) | 0.58 (0.57-0.59) |  |
|  | Age > 69 | 0.67 (0.66-0.68) | 0.43 (0.42-0.44) |  |
|  | Female | 0.77 (0.77-0.77) | 0.58 (0.58-0.58) |  |
|  | Male | 0.70 (0.70-0.70) | 0.48 (0.48-0.48) |  |
|  | 18.5 ≥ BMI < 25 | 0.77 (0.76-0.78) | 0.60 (0.59-0.61) |  |
|  | 25 ≥ BMI < 30 | 0.77 (0.76-0.78) | 0.60 (0.59-0.61) |  |
|  | BMI > 30 | 0.66 (0.66-0.66) | 0.46 (0.46-0.46) |  |
| Fibrosis identification | All | 0.89 (0.88-0.90) | 0.26 (0.24-0.28) | 0.01 |
|  | Asian | 0.72 (0.62-0.82) | 0.14 (0.03-0.25) |  |
|  | Black | 0.90 (0.88-0.92) | 0.22 (0.17-0.27) |  |
|  | White | 0.88 (0.87-0.89) | 0.29 (0.26-0.32) |  |
|  | Other | 0.89 (0.88-0.90) | 0.23 (0.17-0.29) |  |
|  | Age < 40 | 0.83 (0.73-0.93) | 0.06 (0.04-0.08) |  |
|  | Age 40-69 | 0.93 (0.92-0.94) | 0.39 (0.35-0.43) |  |
|  | Age > 69 | 0.83 (0.78-0.88) | 0.20 (0.14-0.26) |  |
|  | Female | 0.87 (0.86-0.88) | 0.26 (0.22-0.30) |  |
|  | Male | 0.89 (0.87-0.91) | 0.27 (0.22-0.32) |  |
|  | 18.5 ≥ BMI < 25 | 0.82 (0.77-0.87) | 0.05 (0.03-0.07) |  |
|  | 25 ≥ BMI < 30 | 0.82 (0.77-0.87) | 0.05 (0.03-0.07) |  |
|  | BMI > 30 | 0.95 (0.94-0.96) | 0.29 (0.26-0.32) |  |

**Abbreviations:** AUROC: area under the precision recall curve. AUPRC: area under the precision recall curve.

**Supplemental Table 16:** Odds ratios per quintile increase in predicted score for MASLD comorbidities among cases

| **Comorbidity** | **UK Biobank** | **MSDW** | **BioMe** | **All of Us** |
| --- | --- | --- | --- | --- |
| MASLD and MetALD | | | | |
| Ischemic heart disease | 1.77 (1.67-1.88) | 1.60 (1.23-2.08) | 1.52 (1.28-1.79) | 1.25 (1.20-1.30) |
| Atrial fibrillation | 1.60 (1.48-1.73) | 1.08 (0.77-1.50) | 1.37 (1.00-1.87) | 1.29 (1.21-1.37) |
| Heart failure | 2.31 (2.01-2.65) | 1.61 (1.02-2.54) | 1.62 (1.20-2.17) | 1.38 (1.30-1.47) |
| Chronic kidney disease | 2.37 (2.12-2.64) | 1.76 (1.16-2.68) | 1.65 (1.32-2.07) | 1.31 (1.24-1.38) |
| Type 2 diabetes | 5.08 (4.63-5.57) | 2.78 (2.19-3.53) | 2.70 (2.33-3.13) | 1.80 (1.72-1.88) |
| Obstructive sleep apnea | 2.49 (2.21-2.79) | 1.32 (1.03-1.71) | 1.54 (1.33-1.77) | 1.38 (1.33-1.44) |
| MASLD only | | | | |
| Ischemic heart disease | 1.83 (1.70-1.97) | 1.55 (1.20-2.00) | 1.56 (1.31-1.85) | 1.22 (1.17-1.28) |
| Atrial fibrillation | 1.57 (1.43-1.73) | 0.98 (0.72-1.34) | 1.30 (0.95-1.79) | 1.27 (1.19-1.36) |
| Heart failure | 2.41 (2.03-2.86) | 1.43 (0.95-2.16) | 1.47 (1.11-1.95) | 1.36 (1.27-1.45) |
| Chronic kidney disease | 2.44 (2.14-2.77) | 1.71 (1.14-2.55) | 1.64 (1.31-2.06) | 1.29 (1.22-1.37) |
| Type 2 diabetes | 5.54 (4.95-6.20) | 2.92 (2.28-3.73) | 2.97 (2.54-3.48) | 1.87 (1.78-1.96) |
| Obstructive sleep apnea | 2.58 (2.25-2.97) | 1.37 (1.06-1.77) | 1.53 (1.32-1.76) | 1.39 (1.33-1.45) |

Values represent odds ratios with 95% confidence intervals.

**Supplemental Table 17:** Odds ratios per quintile increase in predicted score for MASLD comorbidities among controls

| **Comorbidity** | **UK Biobank** | **MSDW** | **BioMe** | **All of Us** |
| --- | --- | --- | --- | --- |
| MASLD and MetALD | | | | |
| Ischemic heart disease | 1.55 (1.54-1.56) | 1.50 (1.47-1.52) | 1.55 (1.51-1.60) | 1.50 (1.48-1.52) |
| Atrial fibrillation | 1.38 (1.36-1.39) | 1.40 (1.37-1.43) | 1.33 (1.29-1.39) | 1.42 (1.39-1.44) |
| Heart failure | 1.76 (1.73-1.79) | 1.67 (1.63-1.71) | 1.69 (1.63-1.75) | 1.83 (1.79-1.88) |
| Chronic kidney disease | 1.88 (1.85-1.91) | 1.93 (1.88-1.98) | 1.71 (1.66-1.76) | 1.71 (1.68-1.74) |
| Type 2 diabetes | 3.86 (3.79-3.92) | 2.37 (2.33-2.42) | 2.82 (2.74-2.91) | 2.58 (2.53-2.62) |
| Obstructive sleep apnea | 2.04 (2.00-2.09) | 1.90 (1.85-1.95) | 1.85 (1.79-1.91) | 1.78 (1.76-1.81) |
| MASLD only | | | | |
| Ischemic heart disease | 1.56 (1.54-1.57) | 1.49 (1.47-1.52) | 1.53 (1.49-1.58) | 1.47 (1.45-1.50) |
| Atrial fibrillation | 1.35 (1.33-1.37) | 1.36 (1.33-1.38) | 1.30 (1.25-1.35) | 1.37 (1.35-1.40) |
| Heart failure | 1.72 (1.69-1.76) | 1.59 (1.56-1.63) | 1.62 (1.57-1.69) | 1.76 (1.72-1.80) |
| Chronic kidney disease | 1.86 (1.83-1.89) | 1.93 (1.88-1.98) | 1.67 (1.62-1.72) | 1.68 (1.65-1.71) |
| Type 2 diabetes | 4.25 (4.16-4.34) | 2.59 (2.53-2.64) | 2.97 (2.88-3.07) | 2.73 (2.68-2.78) |
| Obstructive sleep apnea | 2.16 (2.10-2.22) | 1.88 (1.83-1.93) | 1.83 (1.77-1.90) | 1.79 (1.76-1.81) |

Values represent odds ratios with 95% confidence intervals.

#### Supplemental Table 18: Hazard ratios per quintile increase in predicted score for MASLD outcomes in MSDW

|  | **Cases and controls** | | **Only cases** | | **Only controls** | |
| --- | --- | --- | --- | --- | --- | --- |
| **Outcome** | **Count** | **HR (95% CI)** | **Count** | **HR (95% CI)** | **Count** | **HR (95% CI)** |
| MASLD and MetALD | | | | | | |
| Compensated cirrhosis | 48458 | 1.10 (1.03-1.16) | 568 | 1.87 (1.13-3.08) | 47890 | 1.06 (0.99-1.12) |
| Decompensated cirrhosis | 48496 | 1.08 (1.02-1.14) | 625 | 1.98 (1.10-3.56) | 47871 | 1.05 (0.99-1.11) |
| Any cirrhosis | 48381 | 1.11 (1.05-1.17) | 566 | 1.91 (1.15-3.15) | 47815 | 1.08 (1.02-1.14) |
| Liver cancer | 48552 | 1.04 (0.97-1.1) | 626 | 1.77 (0.93-3.35) | 47926 | 1.02 (0.96-1.08) |
| Liver transplant | 48588 | 1.01 (0.95-1.08) | 641 | 1.19 (0.56-2.52) | 47947 | 1.01 (0.95-1.07) |
| MASLD only | | | | | | |
| Compensated cirrhosis | 46700 | 1.10 (1.03-1.16) | 548 | 1.76 (1.08-2.88) | 46152 | 1.05 (0.99-1.12) |
| Decompensated cirrhosis | 46741 | 1.08 (1.02-1.14) | 605 | 1.98 (1.10-3.58) | 46136 | 1.05 (0.99-1.11) |
| Any cirrhosis | 46626 | 1.11 (1.05-1.17) | 546 | 1.80 (1.11-2.95) | 46080 | 1.08 (1.02-1.14) |
| Liver cancer | 46792 | 1.04 (0.97-1.10) | 605 | 1.78 (0.93-3.41) | 46187 | 1.02 (0.95-1.08) |
| Liver transplant | 46828 | 1.01 (0.95-1.08) | 620 | 1.20 (0.56-2.57) | 46208 | 1.01 (0.94-1.07) |

“Count” represents the number of participants in each analysis. For each outcome, participants with an outcome prior to their cutoff date were excluded. Values represent hazard ratios with 95% confidence intervals. **Abbreviations:** HR: hazard ratio. CI: confidence interval.

**Supplemental Table 19:** Odds ratios per quintile increase in predicted score for MASLD outcomes among all participants

| **Prior diagnosis** | **UK Biobank** | **MSDW** | **BioMe** | **All of Us** |
| --- | --- | --- | --- | --- |
| MASLD and MetALD | | | | |
| Compensated cirrhosis | 2.13 (1.98-2.30) | 2.95 (2.65-3.28) | 1.96 (1.70-2.26) | 2.18 (2.01-2.37) |
| Decompensated cirrhosis | 1.33 (1.29-1.37) | 1.56 (1.46-1.68) | 1.26 (1.13-1.41) | 1.25 (1.21-1.29) |
| Any cirrhosis | 1.40 (1.36-1.44) | 1.90 (1.79-2.03) | 1.51 (1.37-1.66) | 1.35 (1.30-1.39) |
| Liver cancer | 1.52 (1.38-1.68) | 2.11 (1.86-2.39) | 1.50 (1.14-1.99) | 1.57 (1.35-1.81) |
| Liver transplant | 2.14 (1.56-2.93) | 2.10 (1.63-2.70) | 1.80 (1.42-2.29) | 1.65 (1.35-2.03) |
| MASLD only | | | | |
| Compensated cirrhosis | 2.17 (1.98-2.37) | 2.91 (2.61-3.24) | 2.06 (1.77-2.38) | 2.26 (2.07-2.46) |
| Decompensated cirrhosis | 1.33 (1.29-1.38) | 1.57 (1.47-1.69) | 1.26 (1.12-1.41) | 1.24 (1.20-1.29) |
| Any cirrhosis | 1.41 (1.36-1.45) | 1.91 (1.80-2.04) | 1.50 (1.37-1.66) | 1.35 (1.30-1.39) |
| Liver cancer | 1.47 (1.32-1.64) | 2.08 (1.84-2.36) | 1.65 (1.22-2.24) | 1.57 (1.35-1.83) |
| Liver transplant | 2.23 (1.53-3.24) | 2.19 (1.69-2.82) | 1.86 (1.46-2.36) | 1.62 (1.32-1.99) |

**Supplemental Table 20:** Odds ratios per quintile increase in predicted score for MASLD outcomes among cases

| **Prior diagnosis** | **UK Biobank** | **MSDW** | **BioMe** | **All of Us** |
| --- | --- | --- | --- | --- |
| MASLD and MetALD | | | | |
| Compensated cirrhosis | 4.13 (2.98-5.74) | 3.35 (2.32-4.86) | 2.39 (1.58-3.62) | 2.01 (1.77-2.28) |
| Decompensated cirrhosis | 2.49 (1.93-3.21) | 4.04 (1.98-8.24) | 3.54 (1.39-9.01) | 1.41 (1.25-1.58) |
| Any cirrhosis | 2.70 (2.19-3.32) | 3.27 (2.29-4.68) | 2.33 (1.58-3.43) | 1.55 (1.42-1.69) |
| Liver cancer | 2.65 (1.51-4.65) | 3.21 (1.49-6.89) | N/A | 1.91 (1.27-2.87) |
| Liver transplant | N/A | 0.96 (0.48-1.92) | N/A | 2.49 (1.06-5.84) |
| MASLD only | | | | |
| Compensated cirrhosis | 4.72 (3.11-7.17) | 3.73 (2.52-5.52) | 2.46 (1.60-3.77) | 2.02 (1.78-2.30) |
| Decompensated cirrhosis | 2.74 (1.97-3.80) | 6.69 (2.57-17.38) | 4.59 (1.54-13.69) | 1.45 (1.28-1.64) |
| Any cirrhosis | 2.92 (2.25-3.78) | 3.75 (2.55-5.52) | 2.38 (1.60-3.55) | 1.60 (1.46-1.75) |
| Liver cancer | 2.08 (1.16-3.72) | 6.34 (2.09-19.25) | N/A | 2.06 (1.34-3.17) |
| Liver transplant | N/A | 1.36 (0.59-3.16) | N/A | 6.95 (1.08-44.8) |

#### N/A: not applicable due to insufficient events among the cohort.

**Supplemental Table 21:** Odds ratios per quintile increase in predicted score for MASLD outcomes among controls

| **Prior diagnosis** | **UK Biobank** | **MSDW** | **BioMe** | **All of Us** |
| --- | --- | --- | --- | --- |
| MASLD and MetALD | | | | |
| Compensated cirrhosis | 1.84 (1.70-1.99) | 2.16 (1.93-2.42) | 1.57 (1.34-1.84) | 1.73 (1.57-1.90) |
| Decompensated cirrhosis | 1.31 (1.27-1.35) | 1.41 (1.31-1.52) | 1.19 (1.06-1.34) | 1.23 (1.19-1.28) |
| Any cirrhosis | 1.35 (1.31-1.39) | 1.58 (1.48-1.69) | 1.31 (1.18-1.45) | 1.27 (1.22-1.31) |
| Liver cancer | 1.44 (1.30-1.59) | 1.80 (1.58-2.04) | 1.46 (1.09-1.94) | 1.48 (1.26-1.72) |
| Liver transplant | 1.98 (1.44-2.72) | 2.00 (1.52-2.64) | 1.86 (1.45-2.38) | 1.67 (1.35-2.08) |
| MASLD only | | | | |
| Compensated cirrhosis | 1.83 (1.67-2.01) | 2.10 (1.88-2.36) | 1.66 (1.41-1.95) | 1.73 (1.56-1.92) |
| Decompensated cirrhosis | 1.30 (1.26-1.35) | 1.42 (1.32-1.52) | 1.18 (1.04-1.32) | 1.22 (1.18-1.27) |
| Any cirrhosis | 1.35 (1.30-1.40) | 1.58 (1.48-1.69) | 1.30 (1.17-1.44) | 1.26 (1.21-1.31) |
| Liver cancer | 1.40 (1.24-1.57) | 1.74 (1.54-1.98) | 1.60 (1.18-2.19) | 1.42 (1.22-1.67) |
| Liver transplant | 2.16 (1.46-3.20) | 2.02 (1.53-2.67) | 1.93 (1.50-2.48) | 1.63 (1.31-2.02) |

#### Supplemental Table 22: Hazard ratios per quintile increase in predicted score for all-cause and cause-specific mortality in the UK Biobank

| **Disease** | **ICD-10 codes** | **Events** | **HR (95% CI)** |
| --- | --- | --- | --- |
| MASLD and MetALD | | | |
| All causes | N/A | 28678 of 419851 | 1.26 (1.25-1.27) |
| Other digestive diseases | K00-K66, K71-K72, K75-K99 | 1,478 | 1.11 (1.09-1.14) |
| Cerebrovascular disease | I60-I69 | 2,110 | 1.11 (1.10-1.13) |
| Essential hypertension | I10, I12, I15 | 1,913 | 1.19 (1.17-1.22) |
| Extrahepatic cancer | C00-C21, C23-C97 | 15,182 | 1.22 (1.21-1.24) |
| Diabetes mellitus | E10-E14 | 1,404 | 1.23 (1.20-1.25) |
| Heart disease | I00-I09, I11, I13, I20-I51 | 7,829 | 1.33 (1.31-1.35) |
| MASLD only | | | |
| All causes | N/A | 20135 of 291384 | 1.24 (1.23-1.25) |
| Other digestive diseases | K00-K66, K71-K72, K75-K99 | 1,060 | 1.11 (1.08-1.13) |
| Cerebrovascular disease | I60-I69 | 1,506 | 1.11 (1.09-1.13) |
| Essential hypertension | I10, I12, I15 | 1,366 | 1.19 (1.17-1.22) |
| Extrahepatic cancer | C00-C21, C23-C97 | 10,466 | 1.20 (1.19-1.22) |
| Diabetes mellitus | E10-E14 | 1,126 | 1.26 (1.23-1.29) |
| Heart disease | I00-I09, I11, I13, I20-I51 | 5,556 | 1.33 (1.30-1.35) |

For cause-specific mortality, both primary and secondary causes were considered for each disease. **Abbreviations:** HR: hazard ratio. CI: confidence interval. N/A: not applicable.

#### Supplemental Table 23: Correlations between predicted scores and MASLD biomarkers among all participants

|  | **UK Biobank** | | | **MSDW** | | | **All of Us** | | |
| --- | --- | --- | --- | --- | --- | --- | --- | --- | --- |
|  | **Count** | **β** | **SE** | **Count** | **β** | **SE** | **Count** | **β** | **SE** |
| General inflammatory biomarkers | | | | | | | | | |
| CRP | 419097 | 1.82 | 0.02 | 12623 | 2.42 | 0.11 | 30347 | 1.40 | 0.06 |
| ESR | Not measured | | | 19126 | 18.75 | 0.48 | 32422 | 10.88 | 0.29 |
| Other biomarkers | | | | | | | | | |
| GGT | 419706 | 32.25 | 0.13 | 11990 | 80.55 | 2.99 | 5267 | 61.77 | 5.40 |
| CCR | 418844 | -62.92 | 0.33 | Insufficient measurements | | | Insufficient measurements | | |
| Estradiol | 68447 | -223.52 | 4.22 |  |  |  |  |  |  |

All associations were significant to p < 0.001. **Abbreviations:** SE: standard error. CRP: C-reactive protein. ESR: erythrocyte sedimentation rate. GGT: gamma glutamyltransferase. CCR: creatinine/cystatin C ratio.

#### Supplemental Table 24: Correlations between predicted scores and MASLD biomarkers among cases

|  | **UK Biobank** | | | **MSDW** | | | **All of Us** | | |
| --- | --- | --- | --- | --- | --- | --- | --- | --- | --- |
|  | **Count** | **β** | **SE** | **Count** | **β** | **SE** | **Count** | **β** | **SE** |
| General inflammatory biomarkers | | | | | | | | | |
| CRP | 13039 | 1.29 | 0.09 | 138 | 1.71 | 0.86 | 2854 | 0.53 | 0.21 |
| ESR | Not measured | | | 198 | 17.67 | 3.55 | 2883 | 5.25 | 1.02 |
| Other biomarkers | | | | | | | | | |
| GGT | 13052 | 42.27 | 1.05 | 433 | 77.55 | 17.12 | 1048 | 94.08 | 14.41 |
| CCR | 13026 | -26.11 | 1.32 | Insufficient measurements | | | Insufficient measurements | | |
| Estradiol | 1867 | -142.07 | 18.17 |  |  |  |  |  |  |

All associations were significant to p < 0.001 except for CRP in MSDW (p = 0.04) and All of Us (p = 0.01). **Abbreviations:** SE: standard error. CRP: C-reactive protein. ESR: erythrocyte sedimentation rate. GGT: gamma glutamyltransferase. CCR: creatinine/cystatin C ratio.

#### Supplemental Table 25: Correlations between predicted scores and MASLD biomarkers among controls

|  | **UK Biobank** | | | **MSDW** | | | **All of Us** | | |
| --- | --- | --- | --- | --- | --- | --- | --- | --- | --- |
|  | **Count** | **β** | **SE** | **Count** | **β** | **SE** | **Count** | **β** | **SE** |
| General inflammatory biomarkers | | | | | | | | | |
| CRP | 406084 | 1.83 | 0.02 | 12485 | 2.46 | 0.11 | 27493 | 1.54 | 0.07 |
| ESR | Not measured | | | 18928 | 18.99 | 0.48 | 29539 | 11.73 | 0.31 |
| Other biomarkers | | | | | | | | | |
| GGT | 406680 | 31.61 | 0.13 | 11557 | 80.63 | 3.11 | 4219 | 56.65 | 6.09 |
| CCR | 405818 | -63.85 | 0.35 | Insufficient measurements | | | Insufficient measurements | | |
| Estradiol | 66580 | -227.16 | 4.35 |  |  |  |  |  |  |

All associations were significant to p < 0.001. **Abbreviations:** SE: standard error. CRP: C-reactive protein. ESR: erythrocyte sedimentation rate. GGT: gamma glutamyltransferase. CCR: creatinine/cystatin C ratio.

#### Supplemental Table 26: Chart review of Bio*Me* participants without a MASLD diagnosis

|  | **Participant 1** | | **Participant 2** | | **Participant 3** | |
| --- | --- | --- | --- | --- | --- | --- |
| **Set** | **Score** | **Diagnosis** | **Score** | **Diagnosis** | **Score** | **Diagnosis** |
| 1 | 0.03 | No evidence of MASLD | 1.00 | Probable MASLD | 2.04 | No evidence of MASLD |
| 2 | 0.02 | No evidence of MASLD | 1.00 | No evidence of MASLD | 1.93 | Possible MASLD |
| 3 | 0.02 | No evidence of MASLD | 1.00 | Definite MASLD | 1.96 | No evidence of MASLD |
| 4 | 0.04 | No evidence of MASLD | 1.00 | No evidence of MASLD | 1.98 | Possible MASLD (low confidence) |
| 5 | 0.06 | No evidence of MASLD | 1.00 | No evidence of MASLD | 2.04 | Probable MASLD with fibrosis |
| 6 | 0.08 | No evidence of MASLD | 1.00 | No evidence of MASLD | 1.99 | Possible MASLD with fibrosis (low confidence) |
| 7 | 0.06 | No evidence of MASLD | 1.00 | No evidence of MASLD | 1.96 | No evidence of MASLD |
| 8 | 0.06 | No evidence of MASLD | 1.00 | No evidence of MASLD | 1.93 | Possible MASLD with fibrosis |
| 9 | 0.06 | No evidence of MASLD | 1.00 | Possible MASLD | 2.06 | Definite MASLD with cirrhosis |
| 10 | 0.01 | No evidence of MASLD | 1.01 | No evidence of MASLD | 2.00 | No evidence of MASLD |
| 11 | 0.06 | No evidence of MASLD | 1.01 | No evidence of MASLD | 2.04 | Possible MASLD with fibrosis |
| 12 | 0.07 | Possible MASLD | 0.99 | No evidence of MASLD | 1.96 | Definite MASLD |

**Study design:** Participants are matched by age, gender, and ethnicity. Sets 1-4 are White participants; sets 5-8 are Black participants; sets 9-12 are Hispanic participants. The first two sets for each ethnicity have female participants, while the other two have male participants. Ages ranged from 56 to 68 years. **Diagnosis:** Definite MASLD indicates confirmation of MASLD by imaging (ultrasound, CT, or MRI). Probable and possible MASLD are assigned based on clinician assessment of both free text and structured data for risk factors and laboratory measurements.

#### Supplemental Table 27: Ordinal logistic regression for Bio*Me* chart review results

| **Variable** | **OR (95% CI)** | **p value** |
| --- | --- | --- |
| Predicted MASLD score | 4.99 (1.60-15.55) | 0.006 |
| Age | 0.95 (0.83-1.08) | 0.418 |
| Ethnicity | 1.27 (0.51-3.17) | 0.609 |
| Gender | 0.94 (0.21-4.24) | 0.935 |

**Abbreviations:** OR: odds ratio. CI: confidence interval.

### Supplemental Figures

**Supplemental Figure 1:** Association of predicted MASLD scores with mortality risk in UK Biobank and Bio*Me*

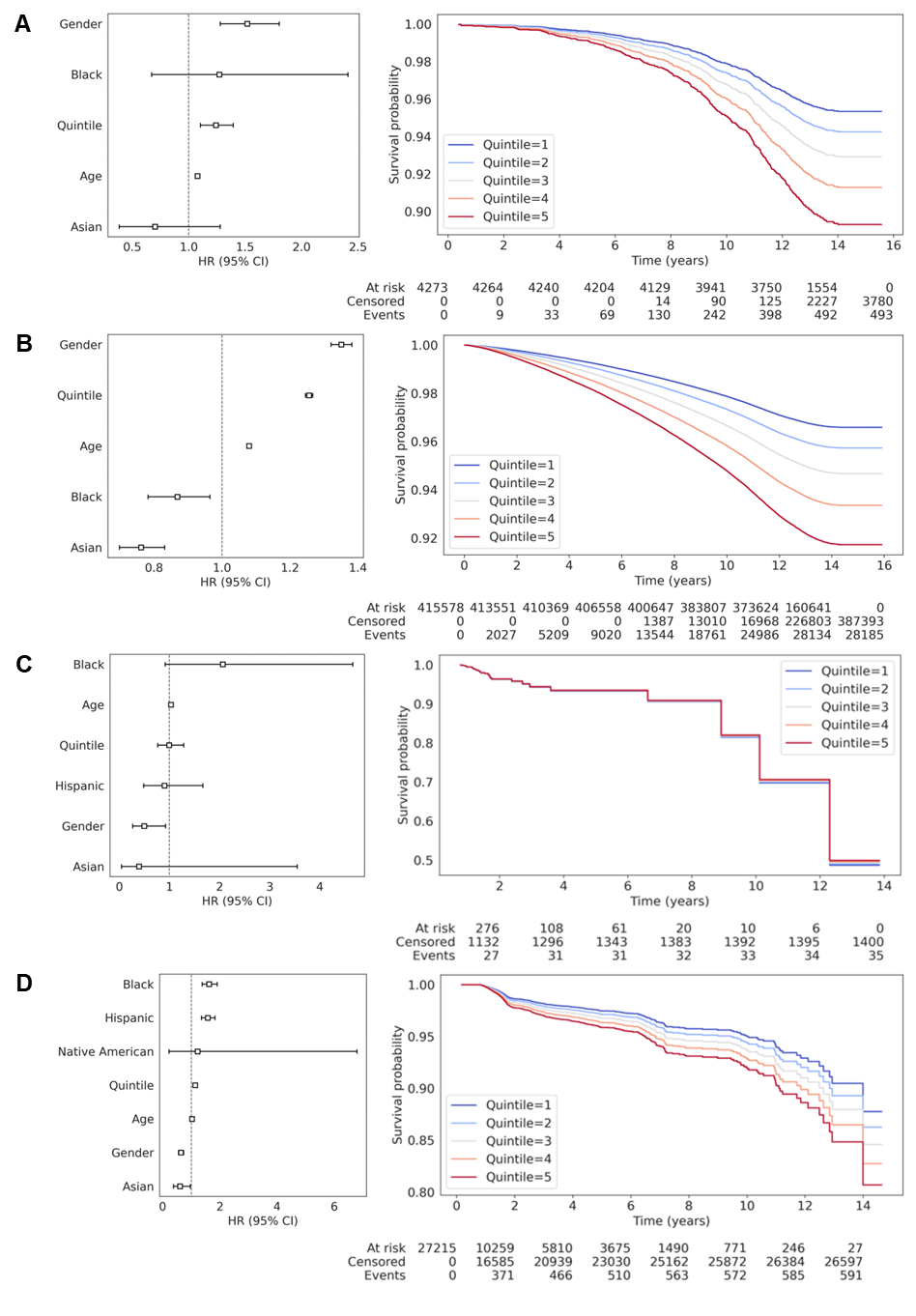

**A-D:** Results of Cox proportional hazards regressions between all-cause mortality and quintile of predicted MASLD score in UK Biobank cases (**A**), UK Biobank controls (**B**), Bio*Me* cases (**C**), and Bio*Me* controls (**D**).
